# Impact of washout duration to account for left truncation in register-based epidemiological studies: a case study on estimating the risk of mental disorders

**DOI:** 10.1101/2025.02.26.25322987

**Authors:** Oleguer Plana-Ripoll, Natalie C. Momen, Dídac Gallego-Alabanda, Danni Chen, Stefan Nygaard Hansen, Carsten Bøcker Pedersen, Esben Agerbo

## Abstract

**Background:** Washout periods can identify and exclude prevalent cases from analyses based on administrative registers with left truncation, but the impact of washout duration on estimates is unknown. We estimated risks of clinically-diagnosed mental disorders according to different washout period durations.

**Methods:** Population-based cohort including all 6,478,162 individuals aged 1-80 years living in Denmark in 2010-2021. Using hospital contacts in 2010-2021, we estimated age-specific incidence rates and cumulative incidence of mental disorders according to different washout period durations (0, 1, 2, 5, 15, and 41 years) based on hospital contacts prior to 2010.

**Results:** Without a washout period, the lifetime cumulative incidence of any mental disorder was 49.4% (95% CI: 49.2%-49.5%) for females and 45.1% (95% CI: 45.0%-45.2%) for males. For each increase in the duration of the washout period to identify and exclude prevalent cases, estimates decreased, reaching a lifetime incidence of 40.3% (95% CI: 40.1%-40.4%) for females and 36.6% (95% CI: 36.5%-36.8%) for males when using all available data (41 years of washout). Without a washout period, estimates for specific mental disorder types were up to 60% higher than those obtained with the maximum washout period, but the bias in absolute terms depended on the underlying risks.

**Conclusions:** Our study is the first to quantify the potential biases related to prevalent cases being incorrectly considered as incident cases. While including all cases identifiable in a register may decrease uncertainty, the inclusion of prevalent cases as being at risk may lead to substantially overestimated measures. We highlight the need for caution when using data sources in which there is left truncation, like administrative registers and electronic healthcare databases.

## Introduction

Administrative registers are a goldmine for epidemiological research because, in some countries, entire populations are included, providing large samples with negligible selection bias.^1,2^ Additionally, information is collected prospectively, minimizing the risk of recall bias.^3^ However, administrative registers have an important limitation that has not received much attention – these registers start collecting data on a specific date, leading to the need to consider left truncation and/or left censoring in analyses.^4^ However, the data to perform such analyses is usually not available in administrative registers as it requires information prior to a register’s inception.

The inception of a register sets the date when information becomes available for administrative and research purposes. For example, information on primary healthcare contacts in Catalonia has been available since 2006,^5^ information on inpatient hospital contacts in Denmark became available in 1969 for psychiatric departments^6^ and in 1977 for all departments,^7^ while the same information has been complete in Sweden since 1987.^8^ The entire lives of individuals born after the inception of the register are captured in the register, to the current date; however, for older individuals, it might be difficult (or impossible) to identify incident cases of chronic conditions if they had onset prior to inception.^7^ For example, someone born in Denmark in 1960 might have received a diagnosis of a mental disorder through a hospital contact before the age of 9 years, but this would not be captured in the register starting in 1969. For individuals born before a register’s inception and having the first record in the register for a specific disease after the inception date, it is unknown whether this first record represents the first hospital contact (i.e., understood as the onset of the disease), or whether it was prevalent, having been diagnosed at an earlier age (see Figure 1 for an illustration).

**Figure 1.**
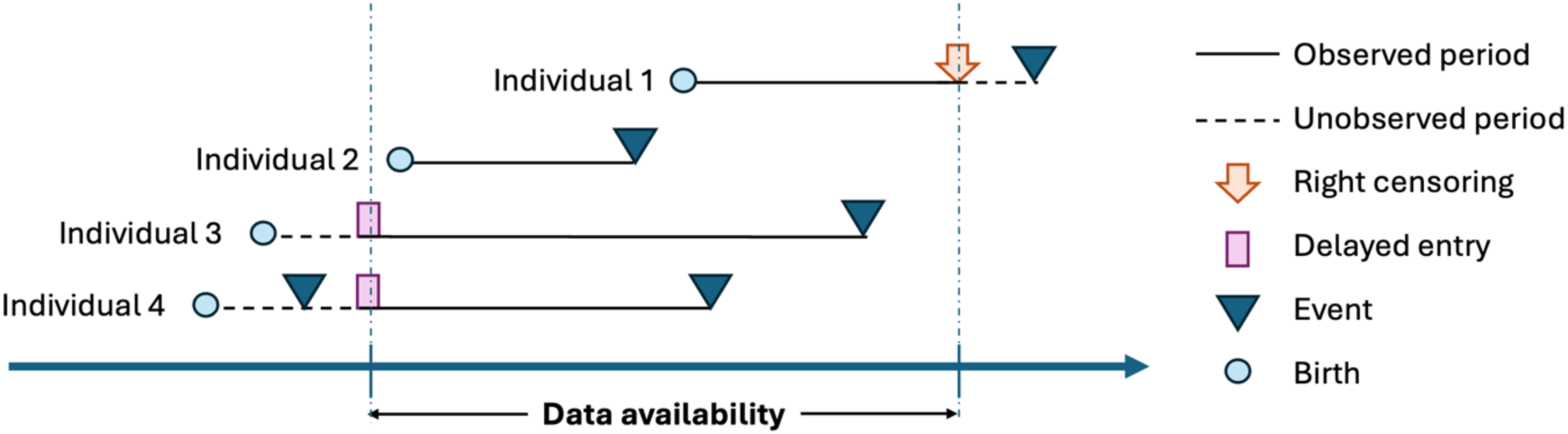
Visualization of left censoring/truncation and right censoring in a hypothetical register-based sample. Individuals 1 and 2 are born after the inception of the register, and their entire lives up to a certain date are captured in the register (individual 1 is right censored without having experienced the event of interest). Individuals 3 and 4 are born before the inception of the register, and they are included in the study with delayed entry when data becomes available. While individual 3 is at risk of developing an incident event, individual 4 should have been identified as a prevalent case. In that case, individual 4 should have been included in the sample as a ‘left censored’ individual or should have been excluded from the sample as a ‘left truncated’ individual. However, the data is not available in the register to identify such case.

In epidemiological studies using traditional patient recruitment, events occurring before baseline may be classified as ‘left censored’ if the individual is enrolled in the study (knowing that the event has happened, perhaps self-reported by the patient, but not knowing when). In that case, special methods are needed to handle left censored data correctly.^4^ Alternatively, data may be considered ‘left truncated’ if individuals with an event before enrolment are disregarded.^9^ This introduces a selection in the sample and means that individuals are included after the *time zero* of interest (i.e., delayed entry) with the knowledge that the event has not occurred prior to their delayed entry into the study.^4^ In other words, risk estimates based on epidemiological follow-up studies should be based on groups of disease-free individuals followed forward for the disease of interest. However, when using administrative registers, only healthcare contacts and not disease onset (from previous contacts) is registered; it is unknown whether individuals are disease-free if they are included with delayed entry. Thus, the first registered contact because of a specific condition in administrative registers may not represent an incident case, but rather a prevalent case. This is particularly likely for older individuals and conditions with onset at young ages.

Measures of disease occurrence, like incidence or prevalence (based on cumulative incidence), may be biased by adding these additional prevalent cases to the set of individuals at risk. In psychiatric epidemiology, administrative registers have been used to estimate age-specific incidence rates and cumulative incidences of mental disorders.^10^ For example, the lifetime cumulative incidence of mental disorders in Denmark is estimated to be 34% for females and 31% for males based on hospital diagnoses,^11^ which surged to an astonishing 87% for females and 77% for males when also including prescriptions for psychotropic medications.^12^ In such studies, no individual is followed over their entire lifespan (due to the more recent inception of available register data), and the standard approach is to use a ‘period survival analysis’,^13,14^ in which different birth cohorts contribute information to different age ranges. Setting a date for the start of follow-up, only those who are at risk after that point are included in the analyses, and survival analysis with delayed entry is used. For example, when the follow-up spans years 2010-2021, individuals born in 1995 will only contribute information to the estimates at ages 15-26 years (Figure 2). For this analysis to provide unbiased estimates, it is necessary that right-censoring is independent, but it is also important to exclude prevalent cases that have been diagnosed before the start of the follow-up. Since registers do not contain information prior to inception, researchers have used ‘washout periods’, which set the start of the follow-up at one specific date at some point *after* the inception of the register and use the hospital contacts in the period between inception and the follow-up start date (i.e., the washout period) to identify and exclude prevalent cases. Previous studies have employed various durations for the washout, e.g., from 1 year of complete data^12^ to 3,^15^ 5^10,16,17^ or even 9 years.^11^ However, no study has investigated the impact of washout duration in identifying prevalent cases, and surprisingly, many studies have only used incomplete data periods (e.g., from inpatient hospitalisations only)^18^ or did not use a washout period at all,^19^ considering all cases in the registers to be incident. Thus, there is a need to quantify the bias introduced by not excluding prevalent cases from the analyses.

**Figure 2.**
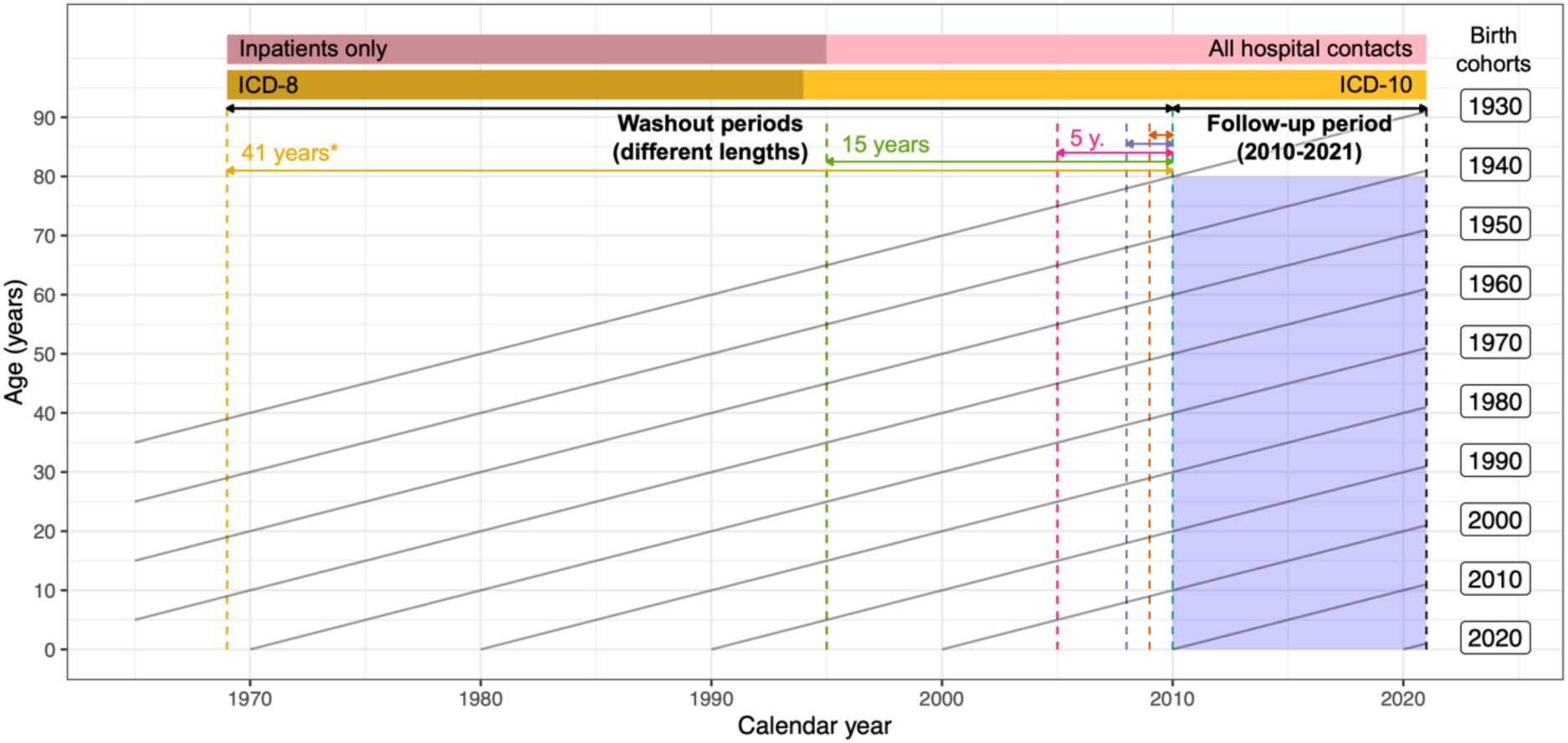
Lexis diagram showing the follow-up period (2010-2021) and different washout periods in relation to the data availability (1969-2009). The diagonal lines indicate individuals belonging to different birth cohorts, linking specific ages during the follow-up with specific calendar years. For example, for individuals born in 1969 (aged 41-52 years during the follow-up period of 2010-2021), the period of registration would cover their entire lives; however, only for those born in 1995 (aged 15-26 years during follow-up) or later would be covered entirely by periods including outpatient contacts and emergency room visits.

The aim of this study is to use complete data from Danish administrative registers to estimate the impact of different durations of washout periods on the age-specific incidence rates and cumulative incidence of mental disorders identified through hospital contacts. Additionally, we provide fully-reproducible R scripts for others to test different washout durations for other types of conditions.

## Methods

### Study population

We designed a population-based cohort study including all 6,478,162 individuals aged 1-80 years living in Denmark at some point between January 1, 2010 and December 31, 2021. Since April 2, 1968, the Danish Civil Registration System^20,21^ has maintained information on all residents, including registered sex (classified as female or male in administrative records), date of birth, continuously updated information on vital status, and a unique personal identification number that is used across all registers.

### Mental disorders

In Denmark, there is free access to health care for the entire population, and hospital contacts are recorded through different registers.^22^ Information on mental disorders was obtained from the Danish Psychiatric Central Research Register,^6^ which contains data on all admissions to psychiatric inpatient facilities since April 1969 (with complete data since 1970) and visits to outpatient psychiatric departments and emergency departments since January 1,1995. The diagnostic system used was the Danish modification of the *International Classification of Diseases, Eighth Revision* (ICD-8) from 1969 to 1993, and *Tenth Revision* (ICD-10) from 1994 onwards. We considered an overall category of any mental disorder (ICD-10 codes F00-F99), as well as several types of mental disorders, diagnosed as inpatient, outpatient, or emergency contacts.^10^ Specific ICD-10 codes are available in Table 1 and corresponding ICD-8 codes are available in Supplementary eTable 1. We only included mental disorders identified through hospital contacts and could not capture milder cases or general mental health issues not requiring a visit with a psychiatrist.

**Table 1.**
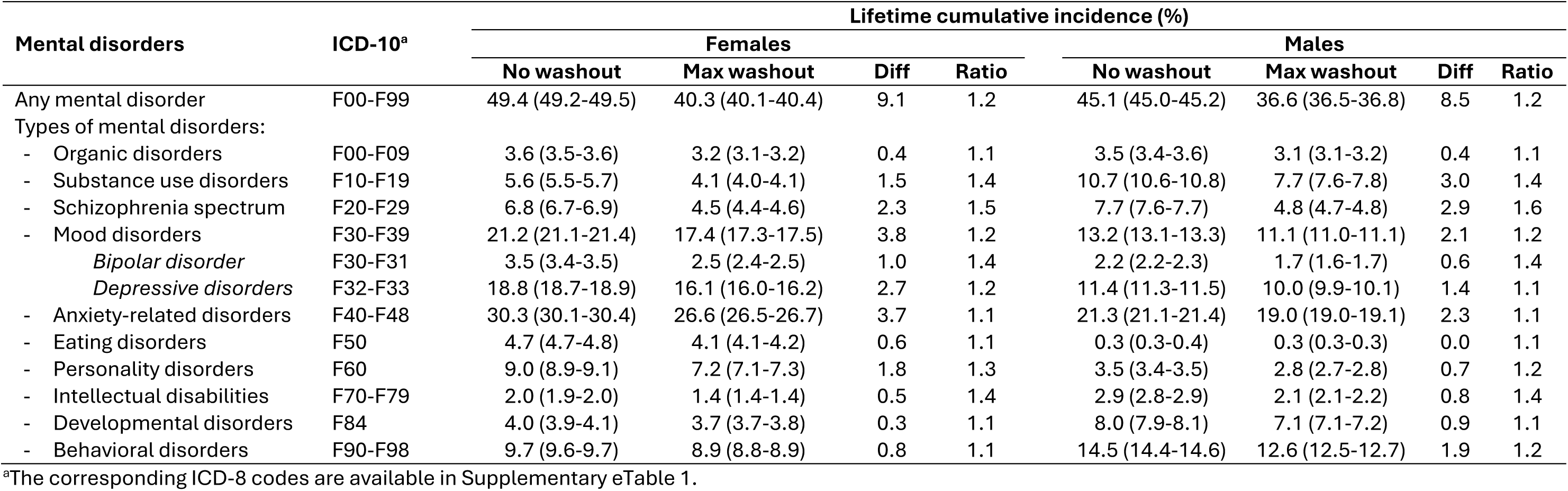
Cumulative incidence of mental disorders at age 80 years for females and males living in Denmark in 2010-2021. Estimates were obtained without using any washout period or using all data available since 1969 (with outpatient data included since 1995). The difference and ratio compare the estimates without a washout period with those using the maximum washout of 41 years. Estimates for each specific washout period (including 1, 2, 5, and 15 years) are available in Supplementary eTable 2.

### Statistical analysis

All analyses were conducted for females and males for each type of disorder using age as time scale. Follow-up for each individual started on January 1, 2010, or at the earliest possible age at which the mental disorder could be developed (Supplementary eTable 1), whichever occurred last. In line with other studies,^10,11,16,17^ diagnoses before the earliest possible age (if any) were ignored. We used different durations of washout periods to identify and exclude prevalent cases. Although we had access to hospital data since 1969, we ignored all data prior to 2010 for the first analysis (0-year washout). Thus, this analysis included the entire population and did not consider any washout period, which would mimic studies that use the entirety of the data period for the follow-up.^18,19^ In the next analysis, we used data from 2009 to exclude prevalent cases, in which the follow-up still started on January 1, 2010, but we excluded all those diagnosed between January 1, 2009 and December 31, 2009 (i.e., 1-year washout) regardless of whether they were also diagnosed after January 1, 2010 or not. In subsequent analyses, we replicated this process using data from 2008 onwards (2-year washout), 2005 onwards (5-year washout), or 1995 onwards (15-year washout). Finally, we performed an analysis using all data available since 1969 (41-year washout), but we note that only inpatient data were captured during the period 1969-1994 and the ICD-10 was implemented in 1994 (ICD-8 was used until 1993); thus, this period was less likely to capture all cases. A Lexis diagram is presented in Figure 2 to demonstrate the concept. All individuals were followed until the date of first diagnosis of any mental disorder (or disorder of interest) recorded in the register, death, emigration from Denmark, or December 31, 2021, whichever occurred first. We estimated age-specific incidence rates and cumulative incidences based on follow-up data in the period 2010-2021 according to the different durations of washout. Age-specific incidence rates were calculated as the number of cases divided by follow-up time according to 5-year age groups, with 95% exact Poisson confidence intervals.^23^ The cumulative incidence of each disorder was estimated using the Aalen-Johansen estimator,^24^ which accounts for the competing risk of dying or emigrating. We considered the lifetime incidence to be the cumulative incidence at age 80 years. All analyses were performed in R Statistical Software (v4.3.2, R core team 2023).

The study was registered with the Danish Data Protection Agency at Aarhus University (No 2016-051-000001-2587) and approved by Statistics Denmark and the Danish Health Data Authority. According to Danish law, informed consent or ethical approval is not required for register-based studies in Denmark.^25^ All data were pseudonymized and not recognizable at an individual level and analyzed on the secure platform of Statistics Denmark. Owing to data protection rules, we are not allowed to share individual-level data. Other researchers who fulfill the requirements set by the data providers could gain access to the data through Statistics Denmark and/or the Danish Health Data Authority. All programming code is available in the supplement.

## Results

A total of 3,241,887 females and 3,236,275 males lived in Denmark at some point between January 1, 2010 and December 31, 2021 while being aged 1-80 years old. During the follow-up period, 271,807 females (8.4%) and 241,652 males (7.5%) were diagnosed with a mental disorder in a psychiatric unit (including outpatient visits).

Considering all these cases as incident cases (i.e., no washout period), the lifetime incidence of any mental disorder was 49.4% (95% CI: 49.2%-49.5%) for females and 45.1% (95% CI: 45.0%-45.2%) for males (Table 1, and Figure 3). However, these estimates decreased when using washout periods to identify and exclude prevalent cases (Supplementary eTable 2), e.g., by 2.7 and 4.5 percentage points for both males and females when using 1 year or 2 years washout periods, respectively, and the estimates decreased even further when using 5 and 15 years, until reaching a lifetime incidence of 40.3% (95% CI: 40.1%-40.4%) for females and 36.6% (95% CI: 36.5%-36.8%) for males when using all data available to identify prevalent cases (41-year washout). Thus, the lifetime cumulative incidence without a washout period was 20% higher than that estimated with the maximum washout period in both females and males, representing a bias in absolute terms of around 9.1 percentage points for females and 8.5 percentage points for males. The age-specific incidence rates of any mental disorder also decreased for each increase in the duration of the washout period, especially for ages 25-65 years, and were more similar for younger and older ages (Figure 3). The number of individuals identified as prevalent, those remaining at risk and number of new cases during follow-up for each duration of washout period is shown in Supplementary eTable 3.

**Figure 3.**
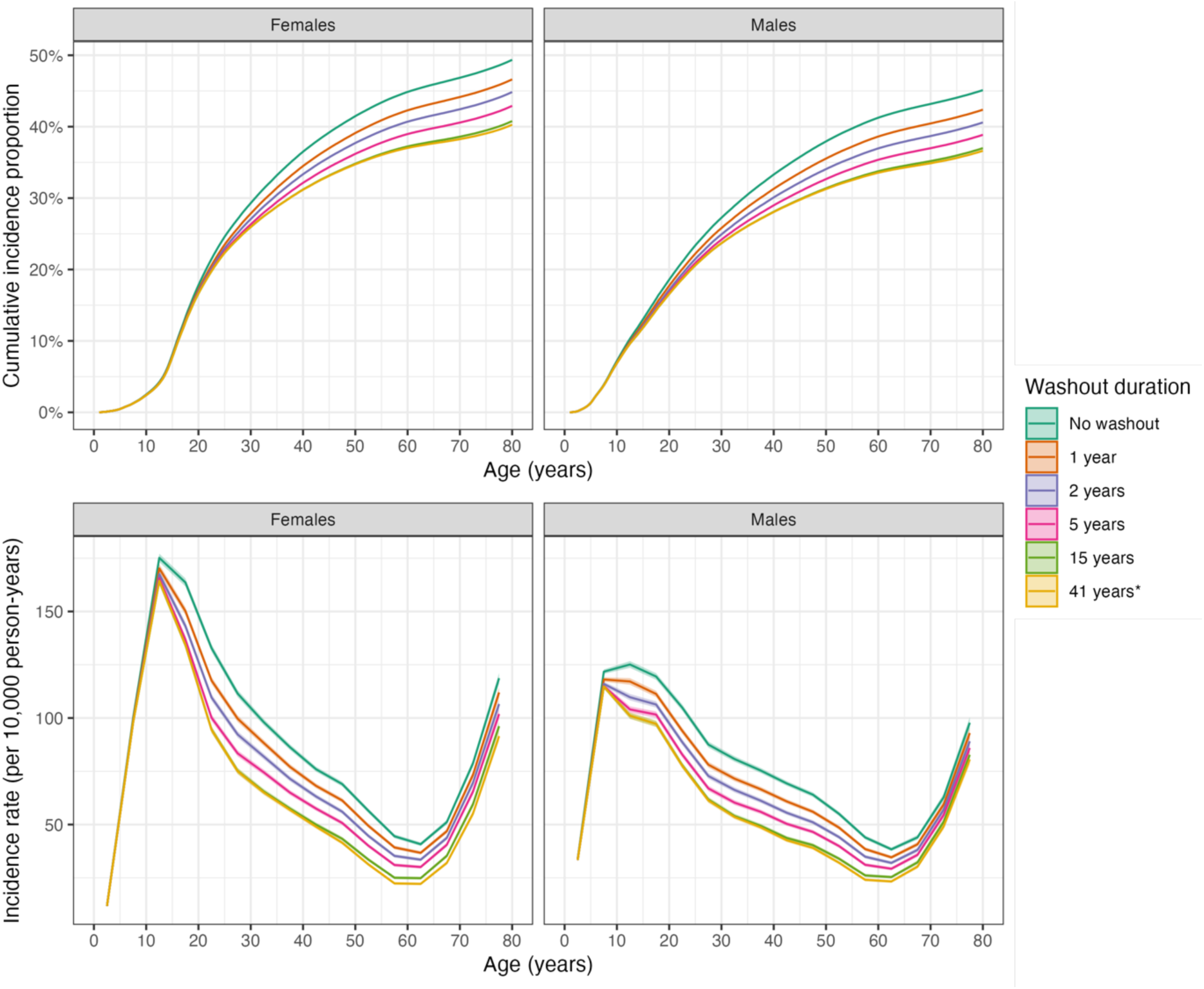
Sex– and age-specific cumulative incidence proportion and incidence rates of any mental disorder in Denmark in 2010-2021 using different durations of washout periods (in the period 1969-2009) to identify and exclude individuals with prevalent diagnoses of mental disorders at start of follow-up.

When looking at specific types of mental disorders, the estimates without a washout period were up to 60% higher than those obtained with the maximum washout period, but the bias in absolute terms depended on the baseline absolute risks (Table 1 and Supplementary eTable 2). For example, we observed similar patterns for prevalent disorders like depressive disorders (Figure 4) and anxiety-related disorders (Supplementary eFigure 1), with around 10-15% higher estimates in relative terms, representing a difference in percentage points of 2.7 and 3.7 for females and 1.4 and 2.3 for males with depressive disorders and anxiety-related disorders, respectively. Results for the overall diagnostic group of mood disorders showed similar patterns (Supplementary eFigure 2). Interestingly, the bias for schizophrenia spectrum disorders was similar in absolute magnitude (2.3 percentage points in females and 2.9 in males), but this represented around 50-60% relative increase due to the lower underlying incidence of the disorder (Figure 5). For substance use disorders, we also observed a considerable difference between the estimates without washout and those with the maximum washout, with a relative increase of around 40% (Figure 6). For all types of mental disorders, the largest differences in age-specific incidence rates when using or not using a washout period were observed for ages 25-65 years, and were more similar for younger and older ages, which explains why the differences in estimates were smaller for disorders occurring during childhood (developmental disorders and behavioral disorders, Supplementary eFigures 3-4) and late adulthood (organic disorders, Supplementary eFigure 5). Estimates for bipolar disorder, eating disorders, personality disorders, and intellectual disabilities are available in Supplementary eFigures 6-9.

**Figure 4.**
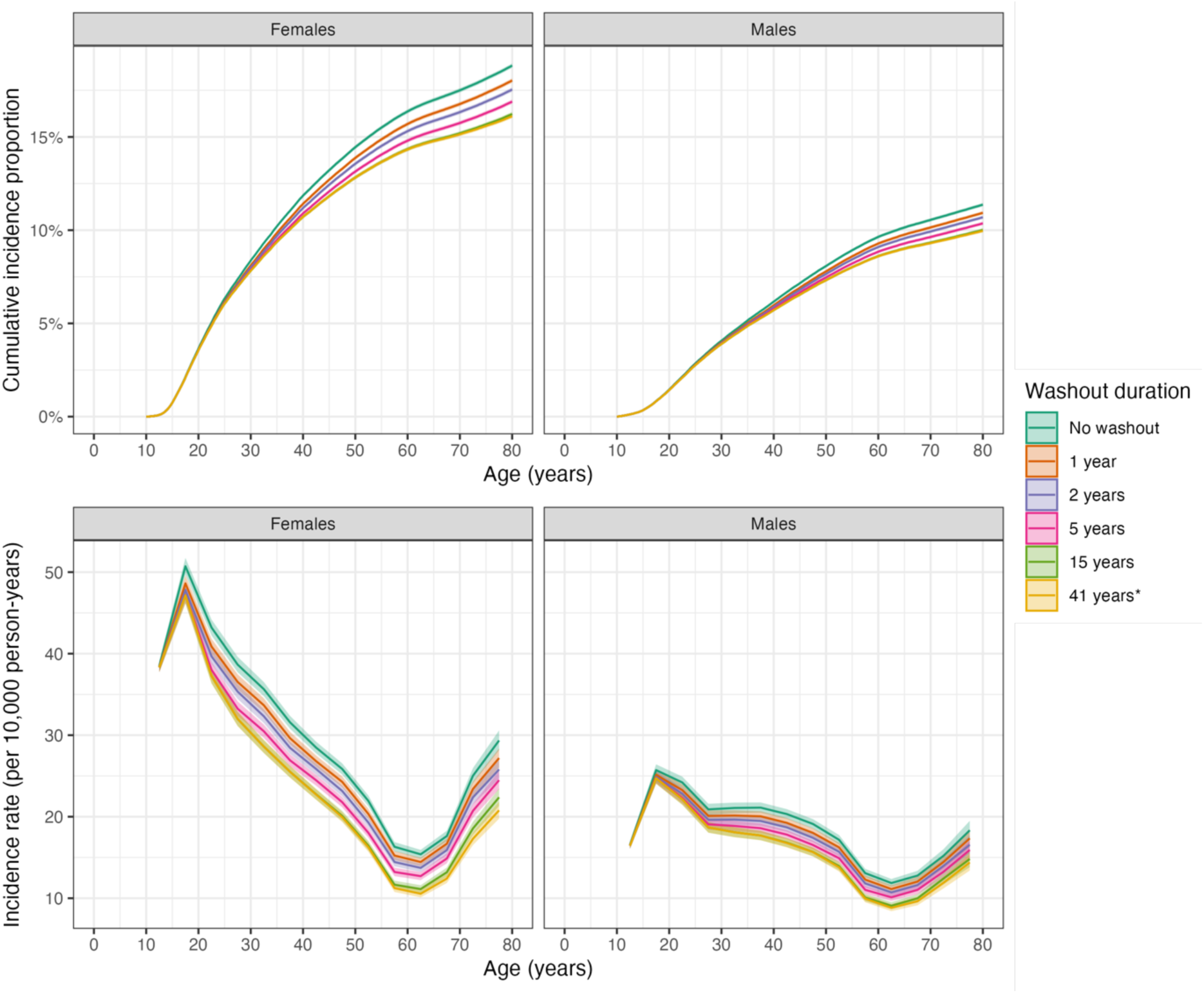
Sex– and age-specific cumulative incidence proportion and incidence rates of depressive disorders in Denmark in 2010-2021 using different durations of washout periods (in the period 1969-2009) to identify and exclude individuals with prevalent diagnoses of depressive disorders at start of follow-up.

**Figure 5.**
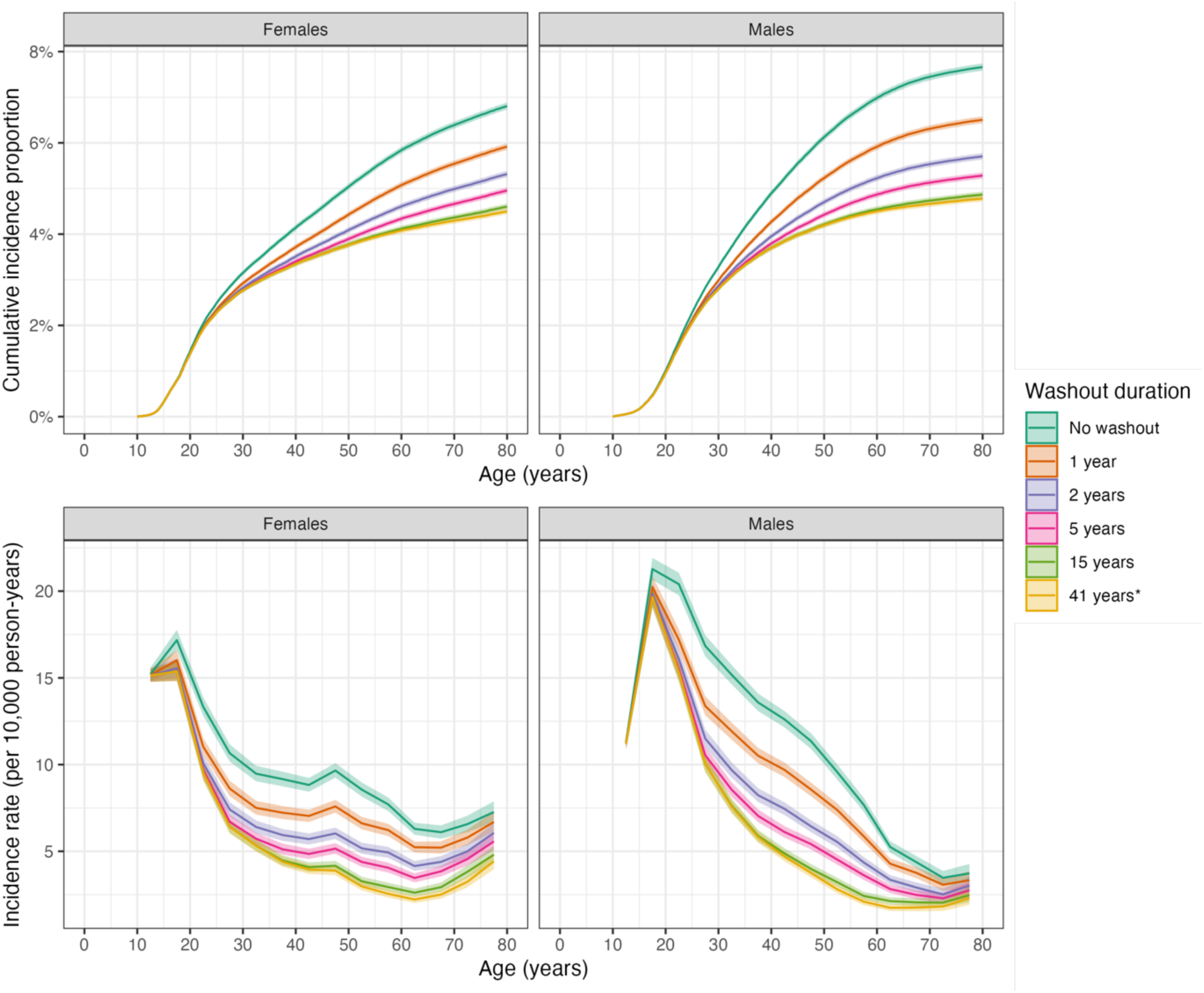
Sex– and age-specific cumulative incidence proportion and incidence rates of schizophrenia spectrum disorders in Denmark in 2010-2021 using different durations of washout periods (in the period 1969-2009) to identify and exclude individuals with prevalent diagnoses of schizophrenia spectrum disorders at start of follow-up.

**Figure 6.**
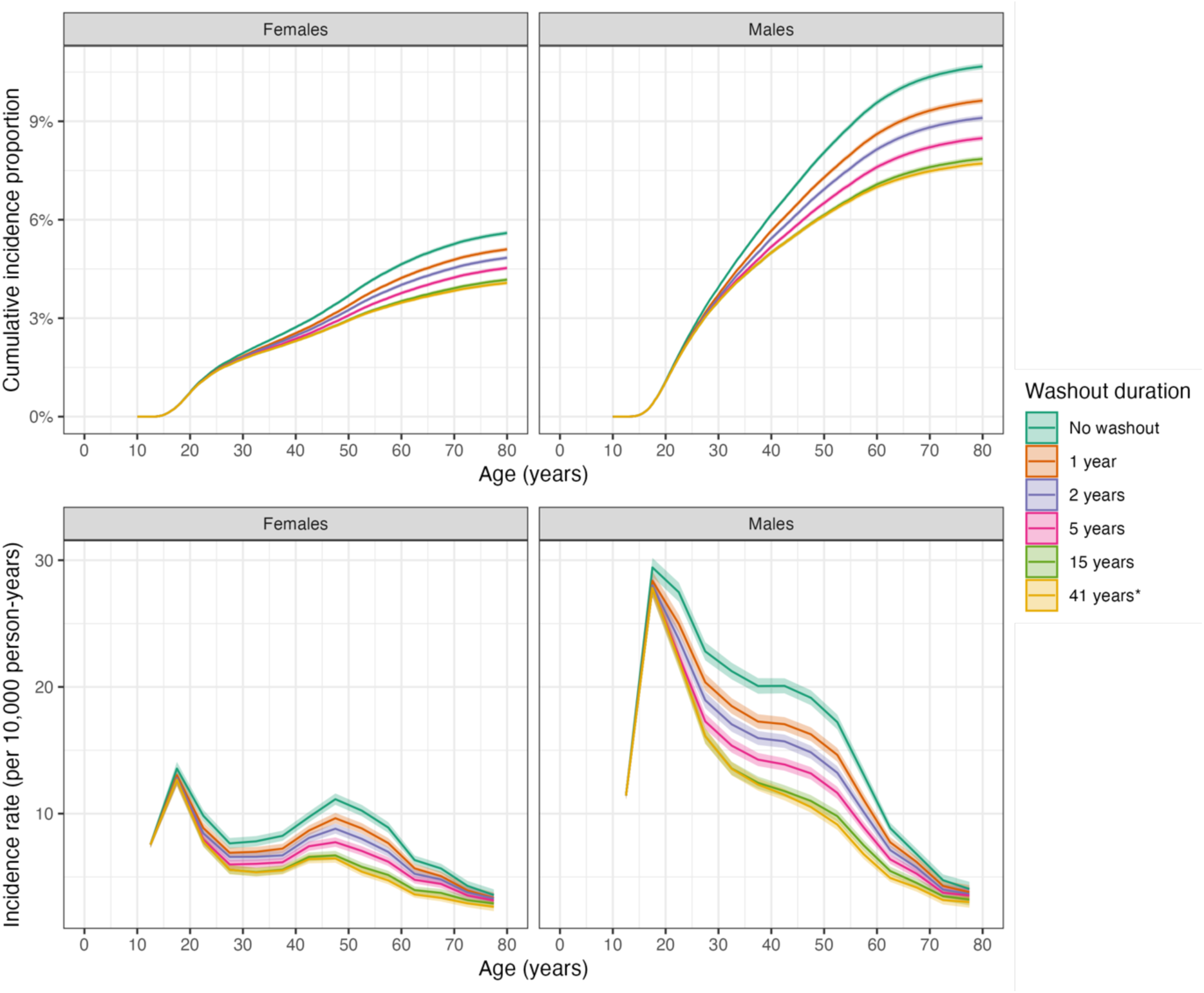
Sex– and age-specific cumulative incidence proportion and incidence rates of substance use disorders in Denmark in 2010-2021 using different durations of washout periods (in the period 1969-2009) to identify and exclude individuals with prevalent diagnoses of substance use disorders at start of follow-up.

## Discussion

We designed this methodological study to investigate the impact of different durations of washout periods on the estimated sex– and age-specific incidence rates and cumulative incidence of mental disorders using Danish national registers. Our results indicated that not employing a washout period resulted in estimates of lifetime cumulative mental disorder incidence that were larger for all included mental disorders in both females and males, compared to employing different durations of washout periods. For females, relative increases in incidence ranged from 50% (for schizophrenia spectrum disorder) to 10% (for organic disorders, anxiety-related disorders, eating disorders, developmental disorders and behavioral disorders). For males, relative increases were between 60% (for schizophrenia spectrum disorder) and 10% (for organic disorders, depressive disorders, anxiety-related disorders, eating disorders, and developmental disorders). For some disorders, these represent substantive absolute differences; for example, for any mental disorder, the absolute difference was 9.1 percentage points for females and 8.5 percentage points for males. The use of age-specific incidence rates in addition to lifetime cumulative incidence allows to explore the bias related to left truncation in short periods for conditions that are episodic in nature, like depression and anxiety.^26^

We observed different patterns in relative and absolute differences according to sex, type of mental disorder, and age group. First, the absolute differences were different for males and females, but the relative differences were very similar. Thus, it is reasonable to assume that sex-differences are explained by different underlying absolute differences in the incidence of mental disorders. Second, the differences between different types of mental disorders might arise from the patterns of disease course trajectories. For example, previous studies have shown that most people diagnosed with schizophrenia in Danish psychiatric hospitals continuously experience symptoms over time,^27^ thereby seeking regular clinical visits; thus, the washout periods (even shorter ones) are more likely to capture prevalent cases. However, individuals treated for depression are less likely to have additional contacts in the following years,^26^ and the differences when using different lengths of washout periods might not be so large for this type of disorders. Third, the largest differences in age-specific incidence when using a washout period compared with not using it were observed for ages 25-65 years, while the differences were smaller for younger and older ages, which might be explained by opposite reasons. At younger ages, even short washout periods are likely to capture prevalent cases; however, those aged 52 years or older do not have their entire lives captured through the period of observation (Figure 2), which implies that, at older ages, even long washout periods are unlikely to capture prevalent cases that occurred at young ages. Thus, at older ages, the cases identified as incident after applying a washout period might still be prevalent cases. An analogous reasoning would explain why the reduction in estimates when using washout periods is not so large for disorders occurring at young ages (e.g., developmental disorders) or at old ages (e.g. organic disorders).

The identification of individuals who are at risk of having an incident disorder is key to obtain valid epidemiological measures in register-based studies. A method based on the ‘waiting time distribution’ – which was graphically presented by Hallas *et al*^28^ and more formally developed by Støvring and Vach^29^ – has been used to estimate incidence and prevalence based on occurrence of individual health service events without knowledge of disease history. This method has been used in pharmacoepidemiology to estimate the proportion of incident and prevalent users among those who redeemed a prescription in a specific time period. However, this method may be more suitable for disorders with frequent contacts with the healthcare services or frequent use of prescribed medications (e.g., insulin^28^ or hypertension^29^), and cannot be used to determine if specific individuals in the sample would be prevalent cases or not (it only allows to estimate population parameters, not individuals’ status). For mental disorders or other conditions with less frequent contacts, the use of washout periods may be the most straightforward method to identify prevalent cases and exclude them from the analyses. Failure to implement a washout period is likely to cause an overestimation of incidence due to misclassification of prevalent cases as incident.^9,30^ This may have been the case in some previous studies, like a recent article in which the authors used a 1-year washout period and reported that the lifetime cumulative incidence of mental disorders and/or prescription of psychotropic medications in Denmark was at around 80% of the population.^12^ In our analyses of mental disorders (described slightly differently and not considering medications), we found that using a 1-year washout period was insufficient to exclude all prevalent cases; the lifetime incidence was reduced by ∼6 percentage points in both females (46.6% to 40.3%) and males (42.4% to 36.6%) when using the maximum duration of washout (41 years) compared with a 1-year washout period only. This issue also translates to health topics other than mental disorders. It could be argued that the impact of a washout period is relatively unproblematic when looking at time trends, as all estimates would be systematically biased. However, it is key that washout periods are applied consistently within different time periods. A recent study by Vinter *et al* used register data to estimate the cumulative incidence of atrial fibrillation and its complications in Denmark in two time periods (2000-2010 and 2011-2022).^31^ The authors used all data available to exclude prevalent cases diagnosed before the start of the follow-up (i.e., they used a washout period). However, the second period (2011-2022) had at least 10 additional years of washout than the first period (2000-2010). Thus, there was more certainty in 2011-2022 than in 2000-2010 that newly identified cases of atrial fibrillation were indeed incident cases, and some of the findings could be explained – at least partly – by a larger overestimation of the estimates in the first period compared to the second one. For studies looking at time trends, it would be important to apply the same duration of washout in all time periods to be sure the overestimation is similar in all periods, even if that meant not using the entire information available. A recent Danish study has applied this approach as a sensitivity analysis to look at time trends in the age-specific incidence rates of mental disorders.^32^

In this study, we highlight potential biases related to not excluding prevalent cases from a sample in presence of left truncation. We have shown that, for mental disorders, the inclusion of prevalent cases as being at risk will lead to substantially overestimated measures for most disorders, in both sexes and age groups. However, it is important to consider that the estimates of cumulative incidence may be severely biased also when there are time trends in the underlying incidence rates, as previously shown by Hansen *et al*.^33^ This is particularly important for mental disorders, which have shown large increases in incidence rates in the recent years.^32,34–36^ The recent increase in incidence might be explained – at least partly – by a ‘catch-up effect’ of seeking mental healthcare for long-term preexisting conditions. In any case, the minimization of bias related to the inclusion of prevalent cases gained through using a washout period might also be affected by underlying time trends, in which shorter washout durations in periods with higher incidence might be equivalent to longer washout durations in periods with lower incidence of mental disorders. When both left truncation and time trends are present, it could be expected that potential overestimation of cumulative incidences would be even greater. Additionally, the inclusion of prevalent cases as being at risk for an incident case overestimates both the number of new cases (numerator in an incidence rate estimator) and the number of individuals at risk (denominator). Thus, although we have shown in this study that the measures of disease occurrence are overestimated for mental disorders, the direction of the bias will depend on the characteristics of the specific condition investigated. Furthermore, the biases in measures of associations caused by including prevalent cases as being at risk for an incident case are unpredictable, particularly when this applies to both an exposed and unexposed group. It could well be that short washout periods are sufficient in these settings if the bias similarly affects those exposed and unexposed; however, investigating these biases is beyond the scope of our current study. To the best of our knowledge, no previous study has investigated the impact of left truncation on measures of association, but this will depend on both the exposure and the outcome of interest.

This study makes use of the large, nationwide registers from Denmark. Data were available for the entire population, thereby minimizing selection bias. Additionally, free and equal access to health care for individuals living in Denmark means that any effects associated with the ability to afford private insurance/access to health care are probably negligible. Additionally, all hospitals must report discharge diagnoses providing data for this study. The generalizability of our findings to other settings (with administrative healthcare data) will depend on many factors, e.g., the coverage and availability of the data. In countries with more private healthcare than in Denmark, it might be necessary to use even longer washout periods. There are several limitations that should be considered when interpreting the results. The analyses only include cases of mental disorders diagnosed within hospital settings; therefore, although those diagnosed in outpatient settings were also included, our findings are not generalizable to milder cases or cases treated in primary care, and we cannot postulate the extent of overestimation without a washout period for cases that would be identified using other data sources within the Danish national register system. For disorders like schizophrenia or bipolar disorder, it is likely all cases have been correctly identified; however, for disorders like depression or anxiety, only the most severe cases would be captured in the hospital register. Thus, the estimates in this study should be interpreted as the incidence of mental disorders diagnosed in hospital settings. We would expect the bias introduced by not considering a washout period to be even larger when all cases of mental disorders would be registered regardless of their severity. Furthermore, the problem with left-truncated data and the identification of prevalent cases is unlikely to be limited to mental disorder research. It is important to also consider the risk of overestimating incidence for diseases other than mental disorders; the R code is made available for other researchers to replicate our findings using other conditions of interest. Finally, despite using different durations of washout periods, we are still not able to include diagnostic information from the years prior to 1969 (or 1995 for outpatients and emergency visits), so the bias is likely still present when using all data available, even if we are able to reduce it.

Employing a washout period can potentially avoid the problem of misclassifying prevalent cases as incident in register-based studies.^10,11,16,17^ In the field of pharmacoepidemiology, researchers using electronic healthcare records have consistently used washout periods to exclude prevalent users from the analyses and apply a new-user design, which is preferable to a prevalent-user design.^37^ However, to the best of our knowledge, our study is the first to quantify the potential biases depending on different washout durations to classify correctly an outcome, in this case mental disorders. While researchers may want to include all cases identifiable in a register, caution needs to be exercised when using data sources where there is left truncation, like in all administrative registers and electronic healthcare databases. It is important to develop new methods that can quantify (and correct for) these biases based on, e.g., plausible assumptions on the distributions of the diseases,^38^ or by extending the waiting-time-distribution method^28,29^ to classify specific individuals and not only estimate population parameters; however, until these methods are easily applicable, register-based epidemiological studies should be carefully designed to avoid misclassifying prevalent cases as incident, which may require using a period of these rich data sources as a washout period to exclude prevalent cases. Researchers should bear in mind that the length of the washout will depend on the nature of the disorder of interest and of the data; for the investigation of acute conditions, one might use only a few weeks of washout if it is relevant to consider the potential repeated periods of the disease over the life course. Nevertheless, it is important to try different lengths of washout periods in sensitivity analyses to explore whether the main results change considerably when increasing the washout duration, which would be an indication of potential prevalent cases being misclassified. Additionally, our findings suggest that washout periods are unlikely to provide additional information for the oldest individuals, for which even the longest washout periods would not cover their childhood and young adulthood. This is particularly important for conditions like mental disorders that tend to occur early in life.^11,39^

## Data Availability

Owing to data protection rules, we are not allowed to share individual-level data. Other researchers who fulfill the requirements set by the data providers could gain access to the data through Statistics Denmark and/or the Danish Health Data Authority.

## SUPPLEMENTARY TABLES AND FIGURES

**eTable 1.** List of disorders included in this study together with the ICD-10 codes and corresponding ICD-8 codes and the minimum possible age of onset for each disorder.

| **Mental disorders** | **ICD-10 codes** | **Corresponding ICD-8 codes** | **Minimum age of onset** |
| --- | --- | --- | --- |
| Any mental disorder | F00-F99 | 290-315 | 1 year |
| Types of mental disorders: |  |  |  |
| - Organic disorders | F00-F09 | 290.09, 290.10, 290.11, 290.18, 290.19, 292.x9, 293.x9, 294.x9 (excluding 294.39), 309.x9 | 35 years |
| - Substance use disorders | F10-F19 | 291.x9, 294.39, 303.x9, 303.20, 303.28, 303.90, 304.x9 | 10 years |
| - Schizophrenia spectrum | F20-F29 | 295.x9, 296.89, 297.x9, 298.29-298.99, 299.04, 299.05, 299.09, 301.83 | 10 years |
| - Mood disorders | F30-F39 | 296.x9 (excluding 296.89), 298.09, 298.19, 300.49, 301.19 | 10 years |
| *Bipolar disorder* | F30-F31 | 296.19, 296.39, 298.19 | 10 years |
| *Depressive disorders* | F32-F33 | 296.09, 296.29, 298.09, 300.49 | 10 years |
| - Anxiety-related disorders | F40-F48 | 300.x9 (excluding 300.49), 305.x9, 305.68, 307.99 | 5 years |
| - Eating disorders | F50 | 306.50, 306.58, 306.59 | 1 year |
| - Personality disorders | F60 | 301.x9 (excluding 301.19), 301.80, 301.81, 301.82, 301.84 | 10 years |
| - Intellectual disabilities | F70-F79 | 311.xx, 312.xx, 313.xx, 314.xx, 315.xx | 1 year |
| - Developmental disorders | F84 | 299.00, 299.01, 299.02, 299.03 | 1 year |
| - Behavioral disorders | F90-F98 | 306.x9 (excluding 306.59), 308.0x | 1 year |

**eTable2.**
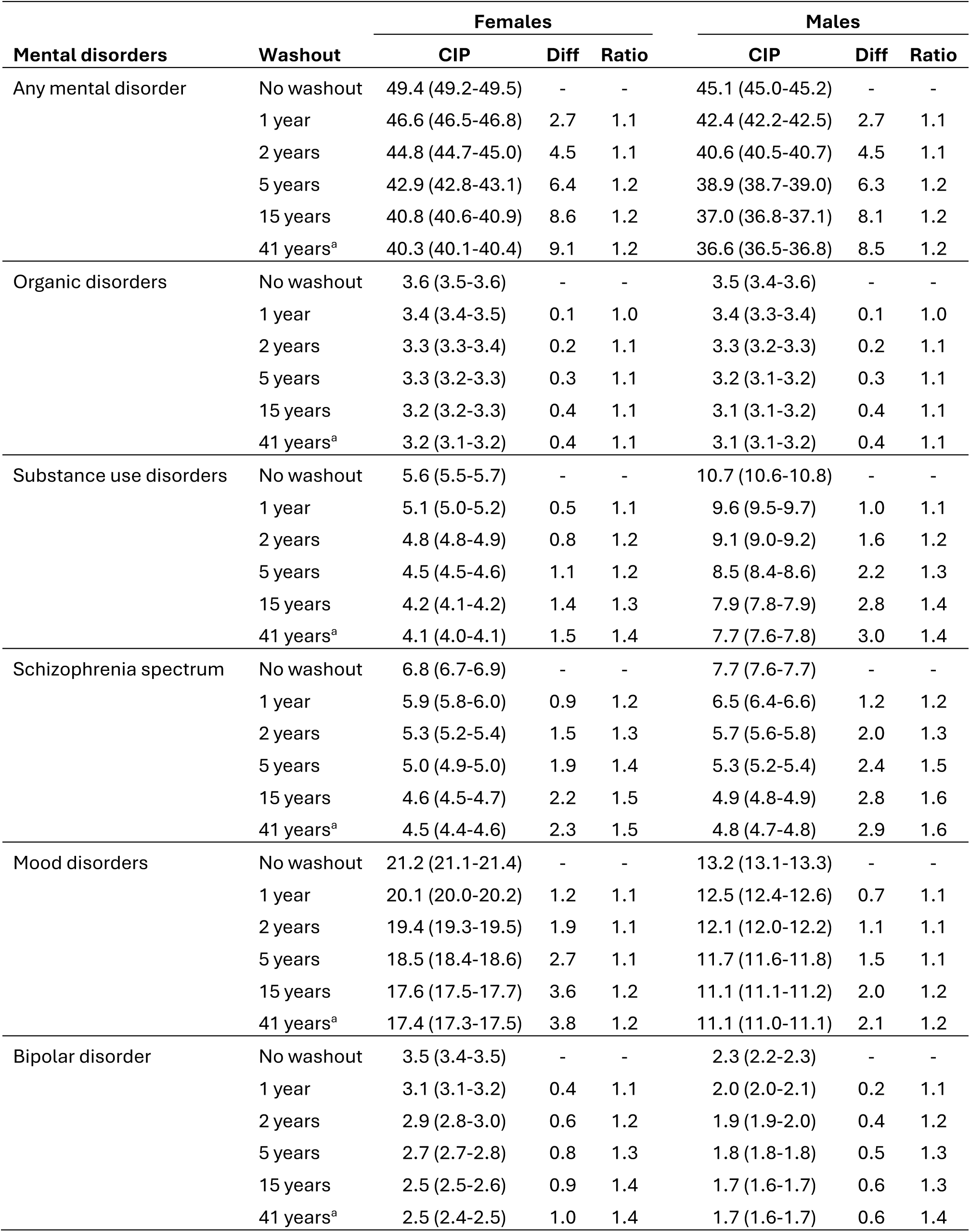
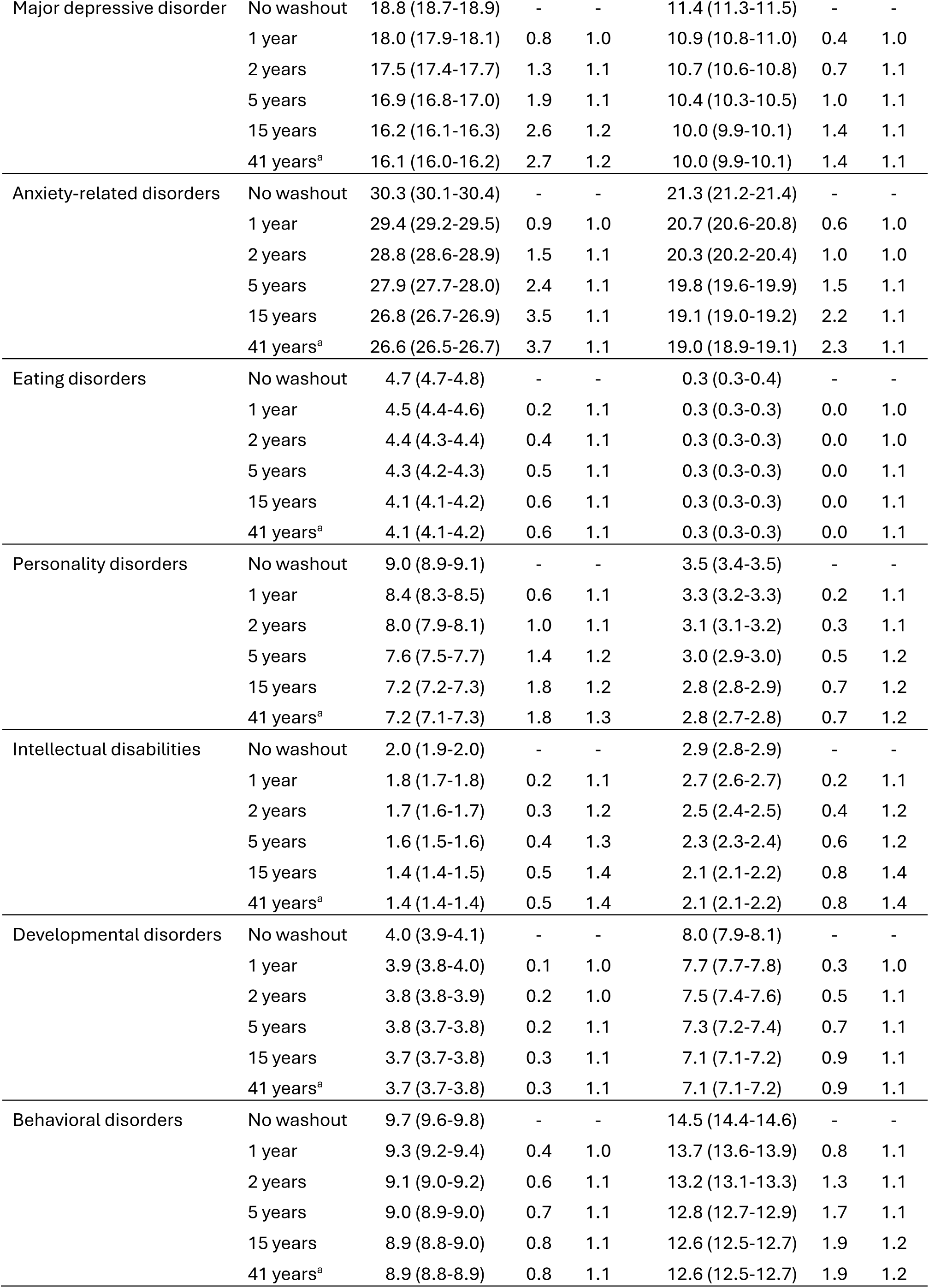
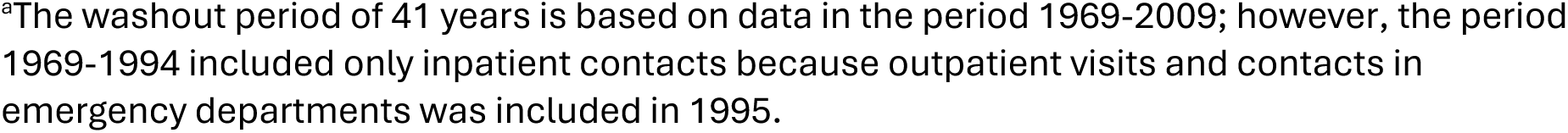
Cumulative incidence proportion (CIP) of mental disorders at age 80 years for females and males living in Denmark in 2010-2021. Estimates were obtained with different washout periods (0, 1, 2, 5, 15 or 41 years, equivalent to using all data available since 1969). The difference and ratio compare the estimates without a washout period with those using the specific washout.

| Mental disorders | Washout | Females |  |  | Males |  |  |
| --- | --- | --- | --- | --- | --- | --- | --- |
|  |  | CIP | Diff | Ratio | CIP | Diff | Ratio |
| Any mental disorder | No washout | 49.4 (49.2-49.5) | - | - | 45.1 (45.0-45.2) | - | - |
|  | 1 year | 46.6 (46.5-46.8) | 2.7 | 1.1 | 42.4 (42.2-42.5) | 2.7 | 1.1 |
|  | 2 years | 44.8 (44.7-45.0) | 4.5 | 1.1 | 40.6 (40.5-40.7) | 4.5 | 1.1 |
|  | 5 years | 42.9 (42.8-43.1) | 6.4 | 1.2 | 38.9 (38.7-39.0) | 6.3 | 1.2 |
|  | 15 years | 40.8 (40.6-40.9) | 8.6 | 1.2 | 37.0 (36.8-37.1) | 8.1 | 1.2 |
|  | 41 years<sup>a</sup> | 40.3 (40.1-40.4) | 9.1 | 1.2 | 36.6 (36.5-36.8) | 8.5 | 1.2 |
| Organic disorders | No washout | 3.6 (3.5-3.6) | - | - | 3.5 (3.4-3.6) | - | - |
|  | 1 year | 3.4 (3.4-3.5) | 0.1 | 1.0 | 3.4 (3.3-3.4) | 0.1 | 1.0 |
|  | 2 years | 3.3 (3.3-3.4) | 0.2 | 1.1 | 3.3 (3.2-3.3) | 0.2 | 1.1 |
|  | 5 years | 3.3 (3.2-3.3) | 0.3 | 1.1 | 3.2 (3.1-3.2) | 0.3 | 1.1 |
|  | 15 years | 3.2 (3.2-3.3) | 0.4 | 1.1 | 3.1 (3.1-3.2) | 0.4 | 1.1 |
|  | 41 years<sup>a</sup> | 3.2 (3.1-3.2) | 0.4 | 1.1 | 3.1 (3.1-3.2) | 0.4 | 1.1 |
| Substance use disorders | No washout | 5.6 (5.5-5.7) | - | - | 10.7 (10.6-10.8) | - | - |
|  | 1 year | 5.1 (5.0-5.2) | 0.5 | 1.1 | 9.6 (9.5-9.7) | 1.0 | 1.1 |
|  | 2 years | 4.8 (4.8-4.9) | 0.8 | 1.2 | 9.1 (9.0-9.2) | 1.6 | 1.2 |
|  | 5 years | 4.5 (4.5-4.6) | 1.1 | 1.2 | 8.5 (8.4-8.6) | 2.2 | 1.3 |
|  | 15 years | 4.2 (4.1-4.2) | 1.4 | 1.3 | 7.9 (7.8-7.9) | 2.8 | 1.4 |
|  | 41 years<sup>a</sup> | 4.1 (4.0-4.1) | 1.5 | 1.4 | 7.7 (7.6-7.8) | 3.0 | 1.4 |
| Schizophrenia spectrum | No washout | 6.8 (6.7-6.9) | - | - | 7.7 (7.6-7.7) | - | - |
|  | 1 year | 5.9 (5.8-6.0) | 0.9 | 1.2 | 6.5 (6.4-6.6) | 1.2 | 1.2 |
|  | 2 years | 5.3 (5.2-5.4) | 1.5 | 1.3 | 5.7 (5.6-5.8) | 2.0 | 1.3 |
|  | 5 years | 5.0 (4.9-5.0) | 1.9 | 1.4 | 5.3 (5.2-5.4) | 2.4 | 1.5 |
|  | 15 years | 4.6 (4.5-4.7) | 2.2 | 1.5 | 4.9 (4.8-4.9) | 2.8 | 1.6 |
|  | 41 years<sup>a</sup> | 4.5 (4.4-4.6) | 2.3 | 1.5 | 4.8 (4.7-4.8) | 2.9 | 1.6 |
| Mood disorders | No washout | 21.2 (21.1-21.4) | - | - | 13.2 (13.1-13.3) | - | - |
|  | 1 year | 20.1 (20.0-20.2) | 1.2 | 1.1 | 12.5 (12.4-12.6) | 0.7 | 1.1 |
|  | 2 years | 19.4 (19.3-19.5) | 1.9 | 1.1 | 12.1 (12.0-12.2) | 1.1 | 1.1 |
|  | 5 years | 18.5 (18.4-18.6) | 2.7 | 1.1 | 11.7 (11.6-11.8) | 1.5 | 1.1 |
|  | 15 years | 17.6 (17.5-17.7) | 3.6 | 1.2 | 11.1 (11.1-11.2) | 2.0 | 1.2 |
|  | 41 years<sup>a</sup> | 17.4 (17.3-17.5) | 3.8 | 1.2 | 11.1 (11.0-11.1) | 2.1 | 1.2 |
| Bipolar disorder | No washout | 3.5 (3.4-3.5) | - | - | 2.3 (2.2-2.3) | - | - |
|  | 1 year | 3.1 (3.1-3.2) | 0.4 | 1.1 | 2.0 (2.0-2.1) | 0.2 | 1.1 |
|  | 2 years | 2.9 (2.8-3.0) | 0.6 | 1.2 | 1.9 (1.9-2.0) | 0.4 | 1.2 |
|  | 5 years | 2.7 (2.7-2.8) | 0.8 | 1.3 | 1.8 (1.8-1.8) | 0.5 | 1.3 |
|  | 15 years | 2.5 (2.5-2.6) | 0.9 | 1.4 | 1.7 (1.6-1.7) | 0.6 | 1.3 |
|  | 41 years<sup>a</sup> | 2.5 (2.4-2.5) | 1.0 | 1.4 | 1.7 (1.6-1.7) | 0.6 | 1.4 |
| Major depressive disorder | No washout | 18.8 (18.7-18.9) | - | - | 11.4 (11.3-11.5) | - | - |
|  | 1 year | 18.0 (17.9-18.1) | 0.8 | 1.0 | 10.9 (10.8-11.0) | 0.4 | 1.0 |
|  | 2 years | 17.5 (17.4-17.7) | 1.3 | 1.1 | 10.7 (10.6-10.8) | 0.7 | 1.1 |
|  | 5 years | 16.9 (16.8-17.0) | 1.9 | 1.1 | 10.4 (10.3-10.5) | 1.0 | 1.1 |
|  | 15 years | 16.2 (16.1-16.3) | 2.6 | 1.2 | 10.0 (9.9-10.1) | 1.4 | 1.1 |
|  | 41 years<sup>a</sup> | 16.1 (16.0-16.2) | 2.7 | 1.2 | 10.0 (9.9-10.1) | 1.4 | 1.1 |
| Anxiety-related disorders | No washout | 30.3 (30.1-30.4) | - | - | 21.3 (21.2-21.4) | - | - |
|  | 1 year | 29.4 (29.2-29.5) | 0.9 | 1.0 | 20.7 (20.6-20.8) | 0.6 | 1.0 |
|  | 2 years | 28.8 (28.6-28.9) | 1.5 | 1.1 | 20.3 (20.2-20.4) | 1.0 | 1.0 |
|  | 5 years | 27.9 (27.7-28.0) | 2.4 | 1.1 | 19.8 (19.6-19.9) | 1.5 | 1.1 |
|  | 15 years | 26.8 (26.7-26.9) | 3.5 | 1.1 | 19.1 (19.0-19.2) | 2.2 | 1.1 |
|  | 41 years<sup>a</sup> | 26.6 (26.5-26.7) | 3.7 | 1.1 | 19.0 (18.9-19.1) | 2.3 | 1.1 |
| Eating disorders | No washout | 4.7 (4.7-4.8) | - | - | 0.3 (0.3-0.4) | - | - |
|  | 1 year | 4.5 (4.4-4.6) | 0.2 | 1.1 | 0.3 (0.3-0.3) | 0.0 | 1.0 |
|  | 2 years | 4.4 (4.3-4.4) | 0.4 | 1.1 | 0.3 (0.3-0.3) | 0.0 | 1.0 |
|  | 5 years | 4.3 (4.2-4.3) | 0.5 | 1.1 | 0.3 (0.3-0.3) | 0.0 | 1.1 |
|  | 15 years | 4.1 (4.1-4.2) | 0.6 | 1.1 | 0.3 (0.3-0.3) | 0.0 | 1.1 |
|  | 41 years<sup>a</sup> | 4.1 (4.1-4.2) | 0.6 | 1.1 | 0.3 (0.3-0.3) | 0.0 | 1.1 |
| Personality disorders | No washout | 9.0 (8.9-9.1) | - | - | 3.5 (3.4-3.5) | - | - |
|  | 1 year | 8.4 (8.3-8.5) | 0.6 | 1.1 | 3.3 (3.2-3.3) | 0.2 | 1.1 |
|  | 2 years | 8.0 (7.9-8.1) | 1.0 | 1.1 | 3.1 (3.1-3.2) | 0.3 | 1.1 |
|  | 5 years | 7.6 (7.5-7.7) | 1.4 | 1.2 | 3.0 (2.9-3.0) | 0.5 | 1.2 |
|  | 15 years | 7.2 (7.2-7.3) | 1.8 | 1.2 | 2.8 (2.8-2.9) | 0.7 | 1.2 |
|  | 41 years<sup>a</sup> | 7.2 (7.1-7.3) | 1.8 | 1.3 | 2.8 (2.7-2.8) | 0.7 | 1.2 |
| Intellectual disabilities | No washout | 2.0 (1.9-2.0) | - | - | 2.9 (2.8-2.9) | - | - |
|  | 1 year | 1.8 (1.7-1.8) | 0.2 | 1.1 | 2.7 (2.6-2.7) | 0.2 | 1.1 |
|  | 2 years | 1.7 (1.6-1.7) | 0.3 | 1.2 | 2.5 (2.4-2.5) | 0.4 | 1.2 |
|  | 5 years | 1.6 (1.5-1.6) | 0.4 | 1.3 | 2.3 (2.3-2.4) | 0.6 | 1.2 |
|  | 15 years | 1.4 (1.4-1.5) | 0.5 | 1.4 | 2.1 (2.1-2.2) | 0.8 | 1.4 |
|  | 41 years<sup>a</sup> | 1.4 (1.4-1.4) | 0.5 | 1.4 | 2.1 (2.1-2.2) | 0.8 | 1.4 |
| Developmental disorders | No washout | 4.0 (3.9-4.1) | - | - | 8.0 (7.9-8.1) | - | - |
|  | 1 year | 3.9 (3.8-4.0) | 0.1 | 1.0 | 7.7 (7.7-7.8) | 0.3 | 1.0 |
|  | 2 years | 3.8 (3.8-3.9) | 0.2 | 1.0 | 7.5 (7.4-7.6) | 0.5 | 1.1 |
|  | 5 years | 3.8 (3.7-3.8) | 0.2 | 1.1 | 7.3 (7.2-7.4) | 0.7 | 1.1 |
|  | 15 years | 3.7 (3.7-3.8) | 0.3 | 1.1 | 7.1 (7.1-7.2) | 0.9 | 1.1 |
|  | 41 years<sup>a</sup> | 3.7 (3.7-3.8) | 0.3 | 1.1 | 7.1 (7.1-7.2) | 0.9 | 1.1 |
| Behavioral disorders | No washout | 9.7 (9.6-9.8) | - | - | 14.5 (14.4-14.6) | - | - |
|  | 1 year | 9.3 (9.2-9.4) | 0.4 | 1.0 | 13.7 (13.6-13.9) | 0.8 | 1.1 |
|  | 2 years | 9.1 (9.0-9.2) | 0.6 | 1.1 | 13.2 (13.1-13.3) | 1.3 | 1.1 |
|  | 5 years | 9.0 (8.9-9.0) | 0.7 | 1.1 | 12.8 (12.7-12.9) | 1.7 | 1.1 |
|  | 15 years | 8.9 (8.8-9.0) | 0.8 | 1.1 | 12.6 (12.5-12.7) | 1.9 | 1.2 |
|  | 41 years<sup>a</sup> | 8.9 (8.8-8.9) | 0.8 | 1.1 | 12.6 (12.5-12.7) | 1.9 | 1.2 |

<sup>a</sup>The washout period of 41 years is based on data in the period 1969-2009; however, the period 1969-1994 included only inpatient contacts because outpatient visits and contacts in emergency departments was included in 1995.

**eTable 3.** For each sex and duration of washout period, number of individuals excluded as being prevalent cases diagnosed with any mental disorder during the washout period, remaining individuals at risk of an incident case, and number of new cases during follow-up.

|  | Washout duration | N | Prevalent cases (excluded) | Individuals at risk | Incident cases during follow-up |
| --- | --- | --- | --- | --- | --- |
| **Females** |  |  |  |  |  |
|  | 0 years | 3,241,887 | 0 | 3,241,887 | 271,807 |
|  | 1 year | 3,241,887 | 34,896 | 3,206,991 | 249,724 |
|  | 2 years | 3,241,887 | 63,050 | 3,178,837 | 235,333 |
|  | 5 years | 3,241,887 | 106,673 | 3,135,214 | 219,363 |
|  | 15 years | 3,241,887 | 180,771 | 3,061,116 | 201,154 |
|  | 41 years<sup>a</sup> | 3,241,887 | 220,383 | 3,021,504 | 195,849 |
| **Males** |  |  |  |  |  |
|  | 0 years | 3,236,275 | 0 | 3,236,275 | 241,652 |
|  | 1 year | 3,236,275 | 31,772 | 3,204,503 | 221,138 |
|  | 2 years | 3,236,275 | 56,123 | 3,180,152 | 207,863 |
|  | 5 years | 3,236,275 | 91,128 | 3,145,147 | 194,677 |
|  | 15 years | 3,236,275 | 150,369 | 3,085,906 | 179,892 |
|  | 41 years<sup>a</sup> | 3,236,275 | 179,358 | 3,056,917 | 176,359 |

<sup>a</sup>The washout period of 41 years is based on data in the period 1969-2009; however, the period 1969-1994 included only inpatient contacts because outpatient visits and contacts in emergency departments was included in 1995.

**eFigure 1.**
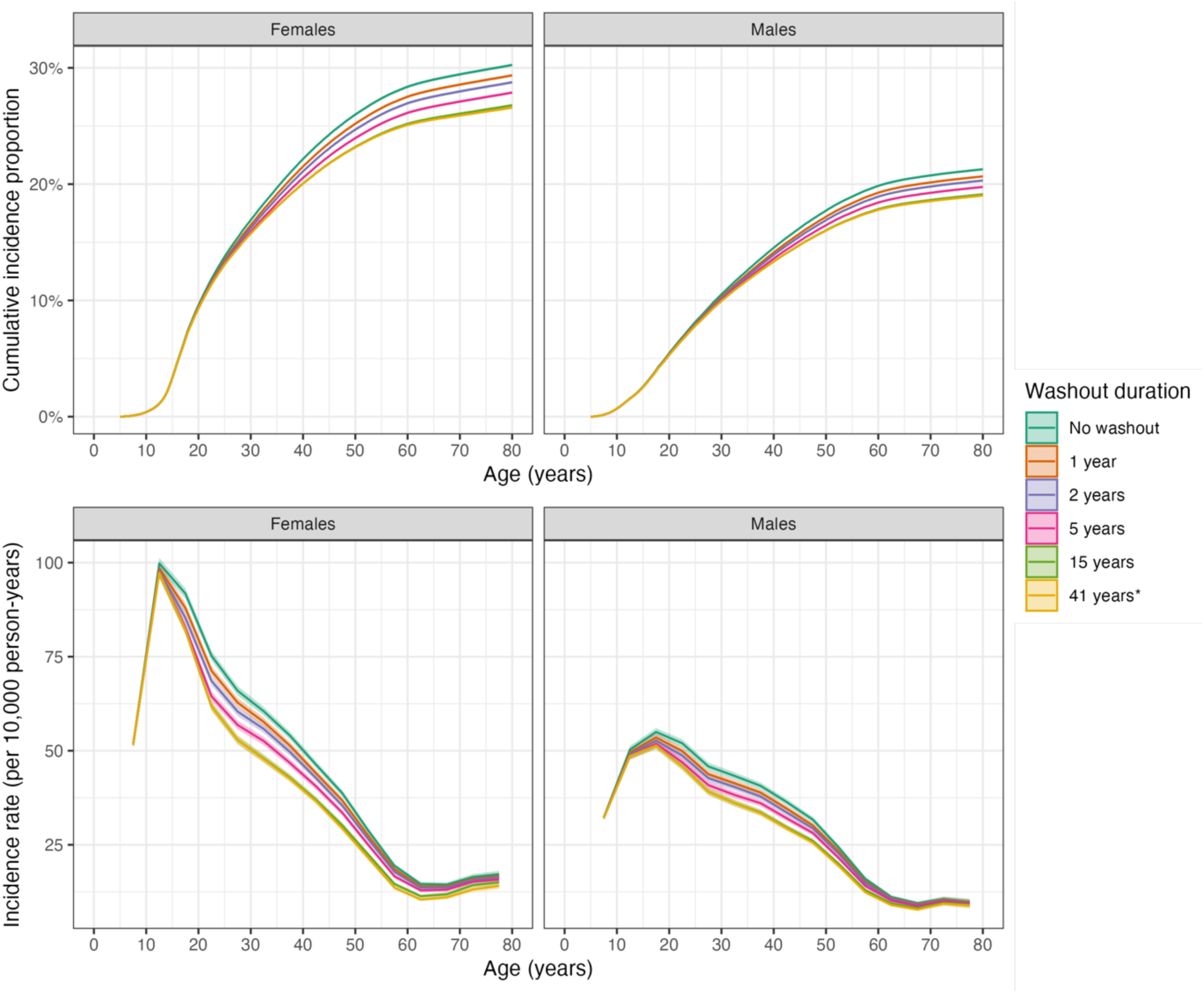
Sex– and age-specific cumulative incidence proportion and incidence rates of anxiety-related disorders in Denmark in 2010-2021 using different durations of washout periods (in the period 1969-2009) to identify and exclude individuals with prevalent diagnoses of anxiety-related disorders at start of follow-up.

**eFigure 2.**
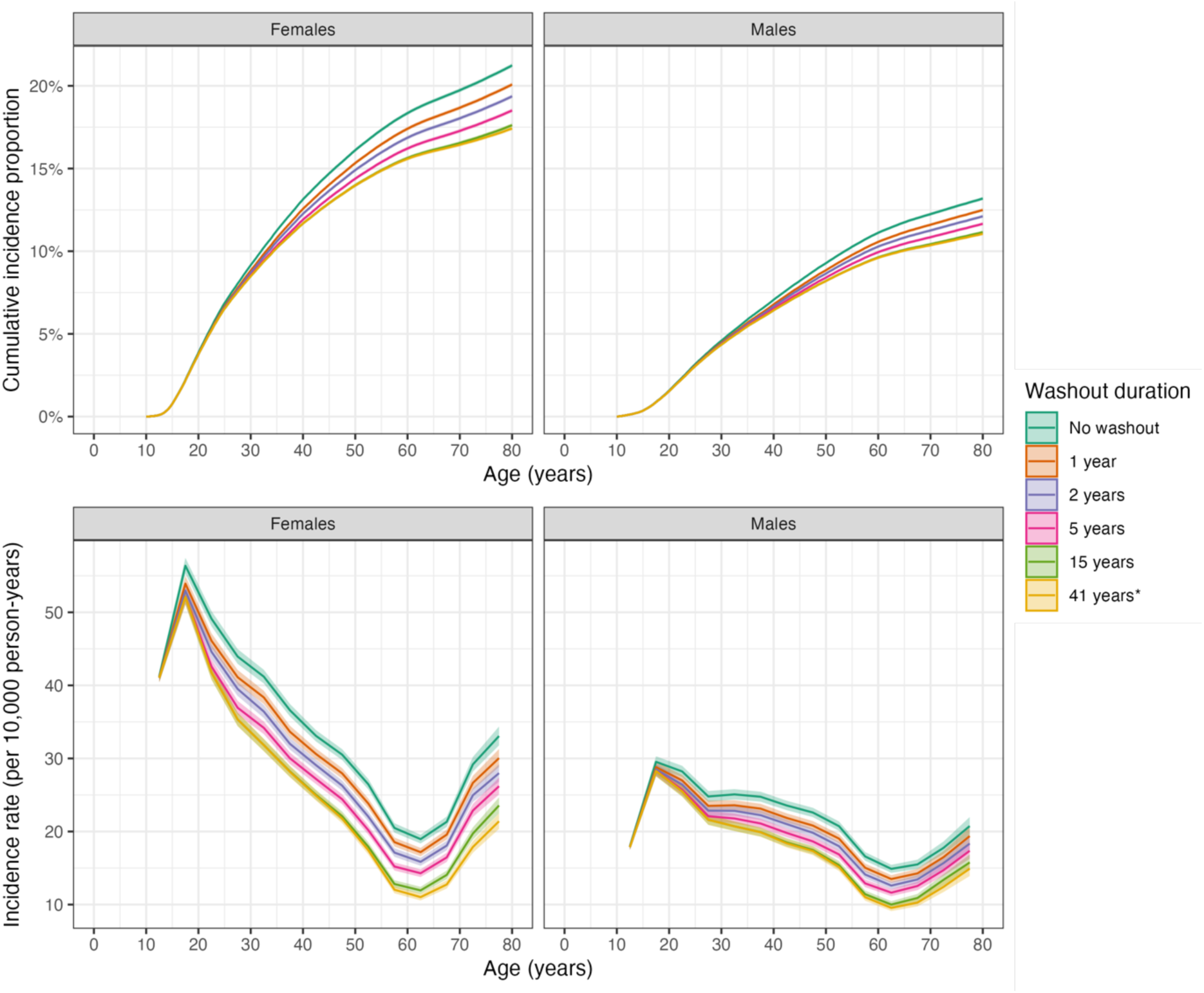
Sex– and age-specific cumulative incidence proportion and incidence rates of mood disorders in Denmark in 2010-2021 using different durations of washout periods (in the period 1969-2009) to identify and exclude individuals with prevalent diagnoses of mood disorders at start of follow-up.

**eFigure 3.**
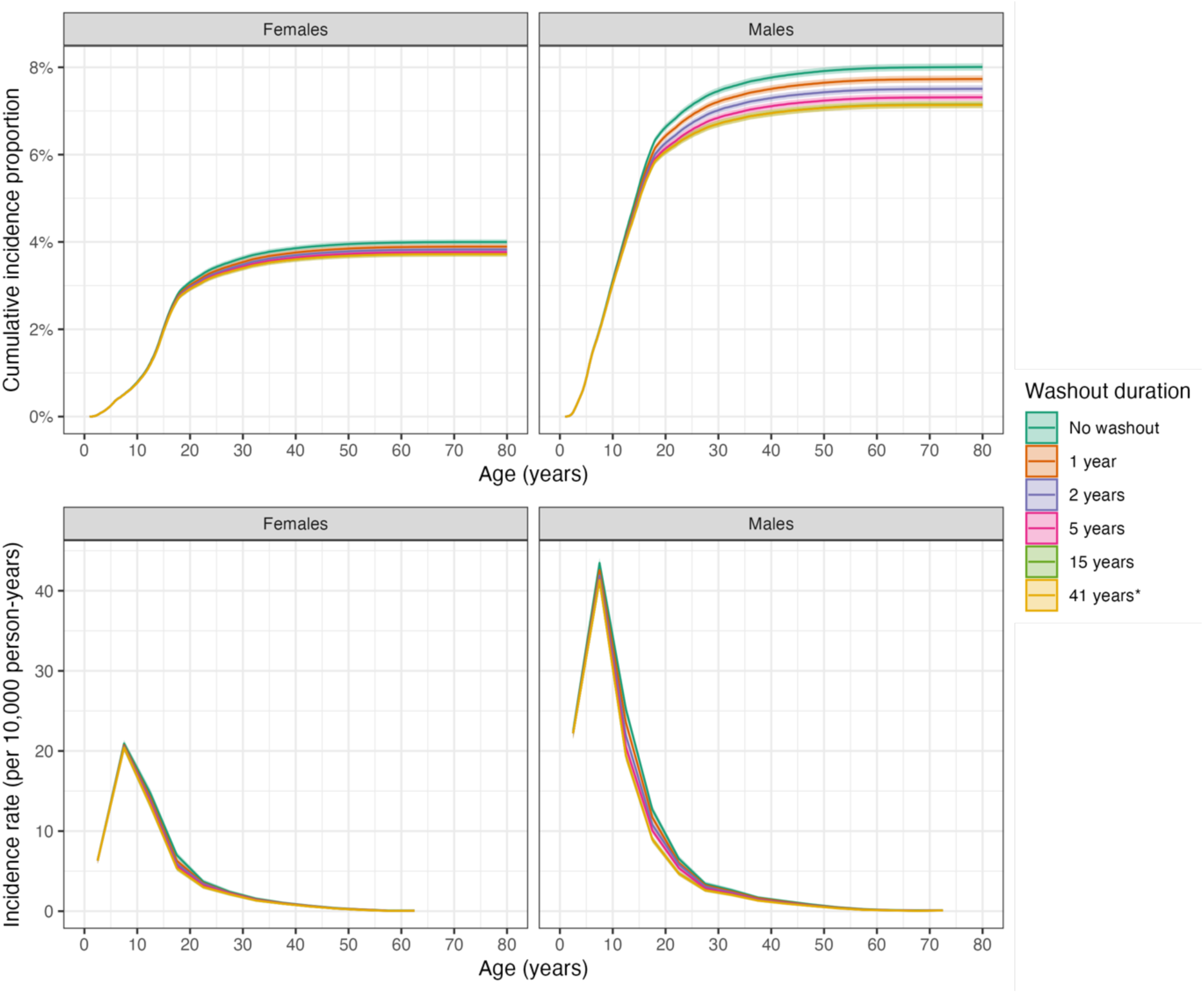
Sex– and age-specific cumulative incidence proportion and incidence rates of developmental disorders in Denmark in 2010-2021 using different durations of washout periods (in the period 1969-2009) to identify and exclude individuals with prevalent diagnoses of developmental disorders at start of follow-up.

**eFigure 4.**
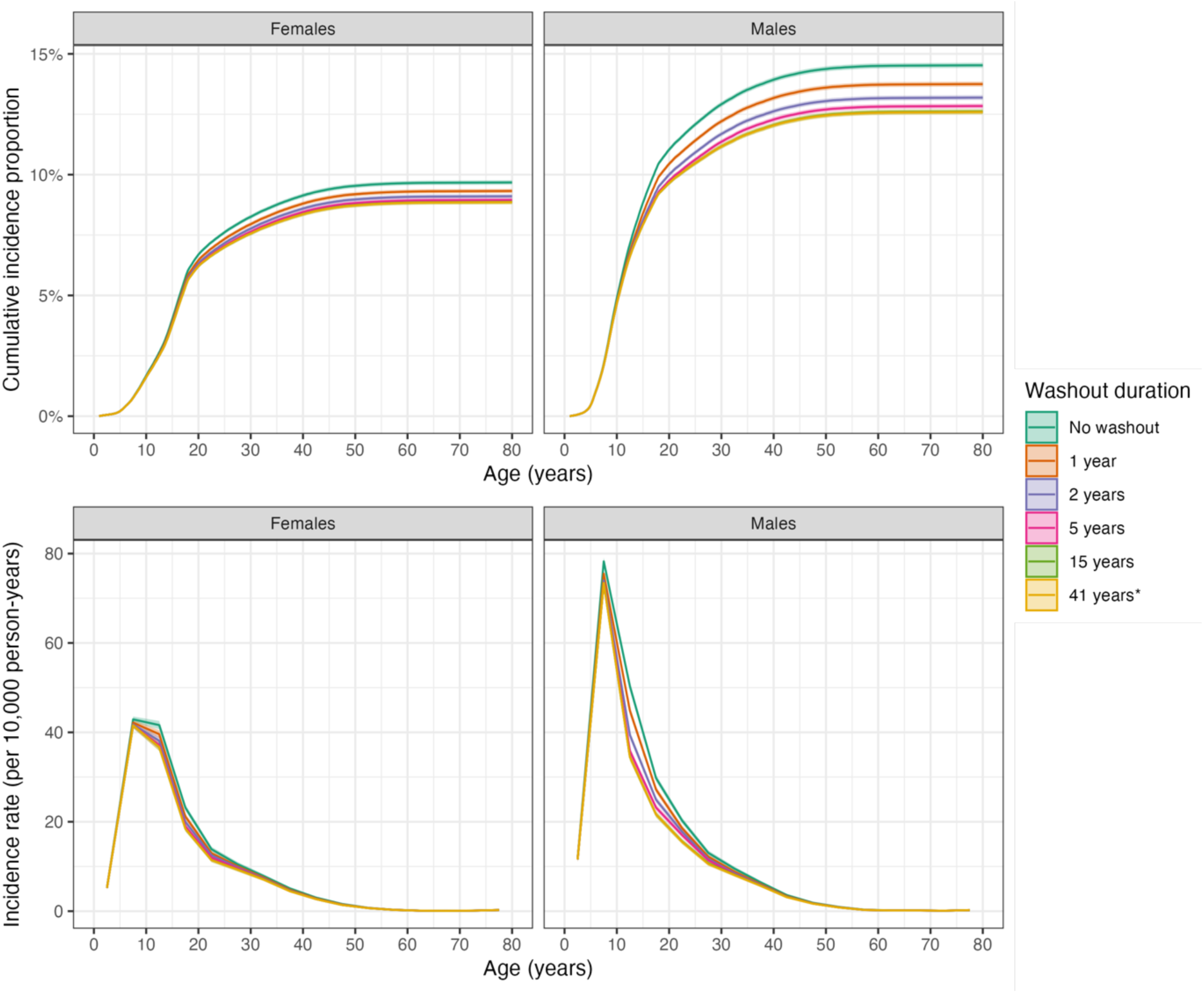
Sex– and age-specific cumulative incidence proportion and incidence rates of behavioral disorders in Denmark in 2010-2021 using different durations of washout periods (in the period 1969-2009) to identify and exclude individuals with prevalent diagnoses of behavioral disorders at start of follow-up.

**eFigure 5.**
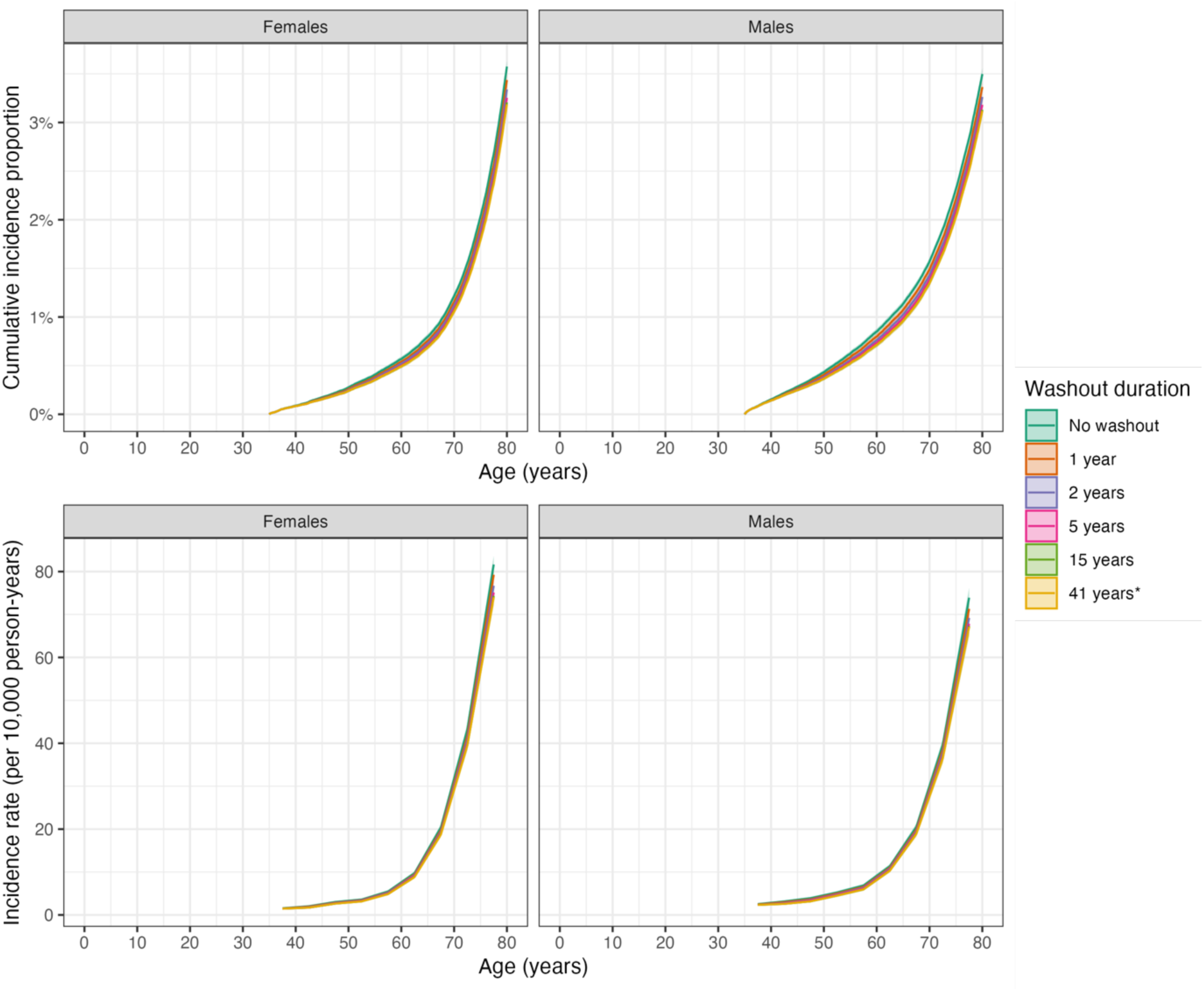
Sex– and age-specific cumulative incidence proportion and incidence rates of organic disorders in Denmark in 2010-2021 using different durations of washout periods (in the period 1969-2009) to identify and exclude individuals with prevalent diagnoses of organic disorders at start of follow-up.

**eFigure 6.**
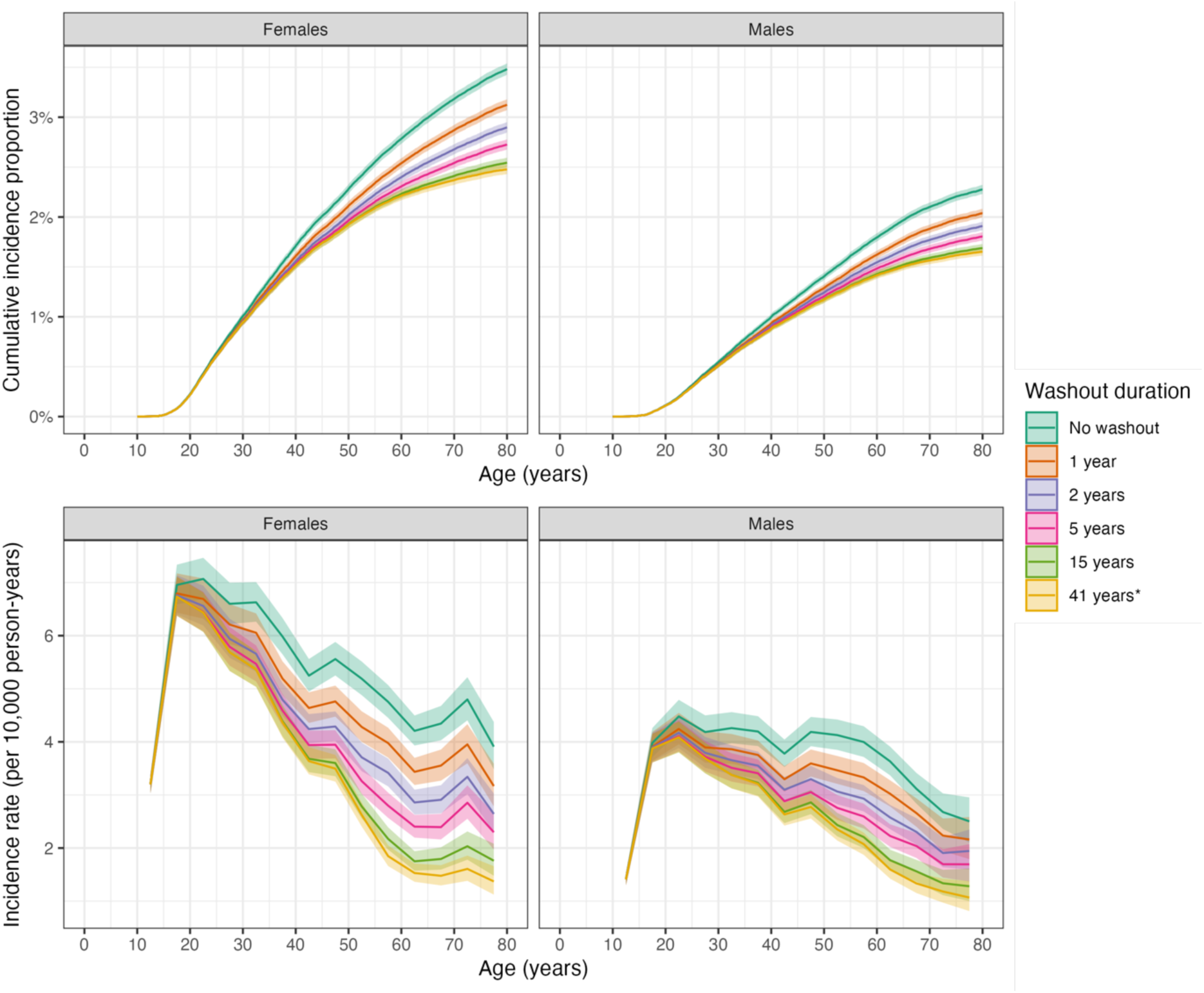
Sex– and age-specific cumulative incidence proportion and incidence rates of bipolar disorder in Denmark in 2010-2021 using different durations of washout periods (in the period 1969-2009) to identify and exclude individuals with prevalent diagnoses of bipolar disorder at start of follow-up.

**eFigure 7.**
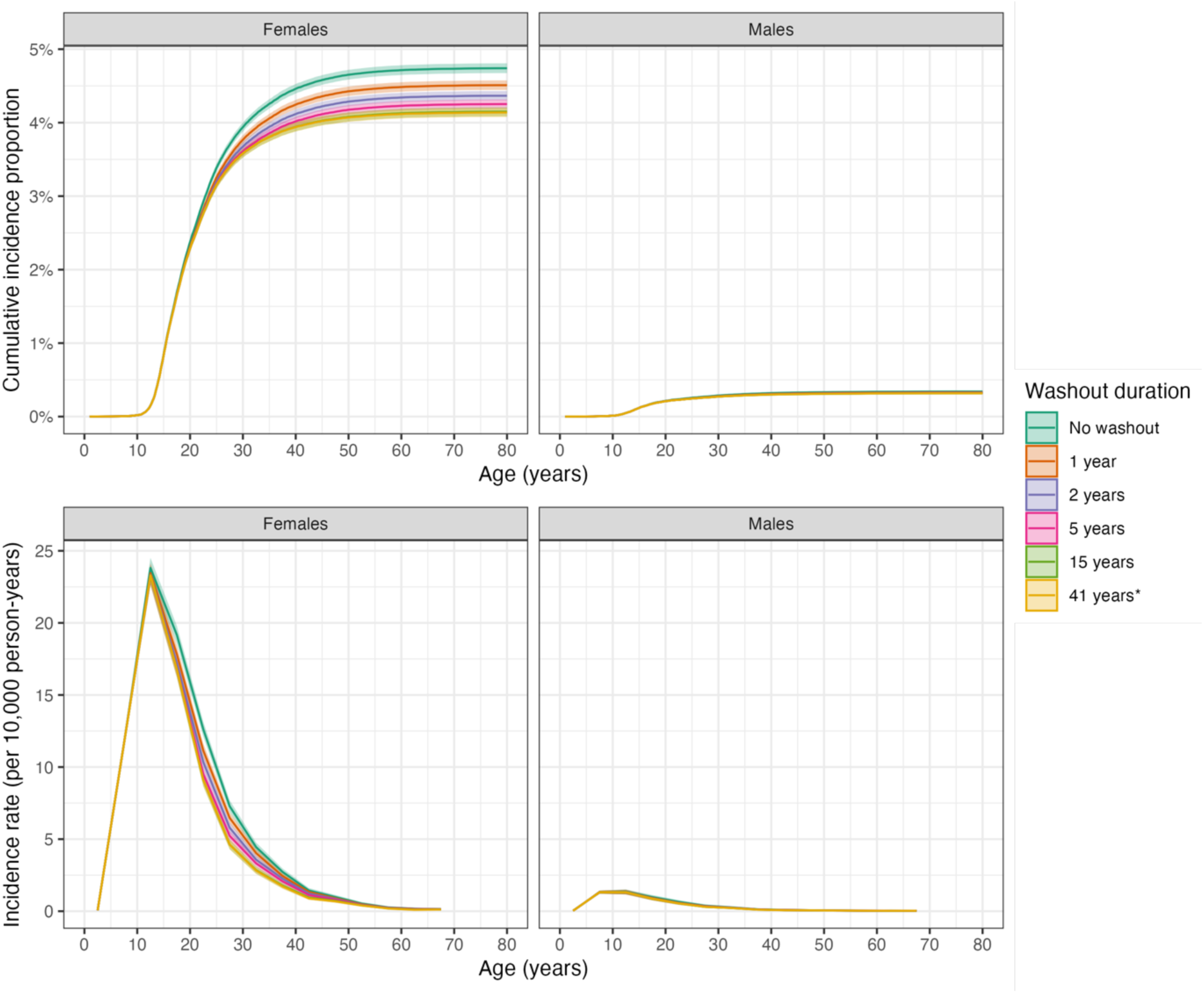
Sex– and age-specific cumulative incidence proportion and incidence rates of eating disorders in Denmark in 2010-2021 using different durations of washout periods (in the period 1969-2009) to identify and exclude individuals with prevalent diagnoses of eating disorders at start of follow-up.

**eFigure 8.**
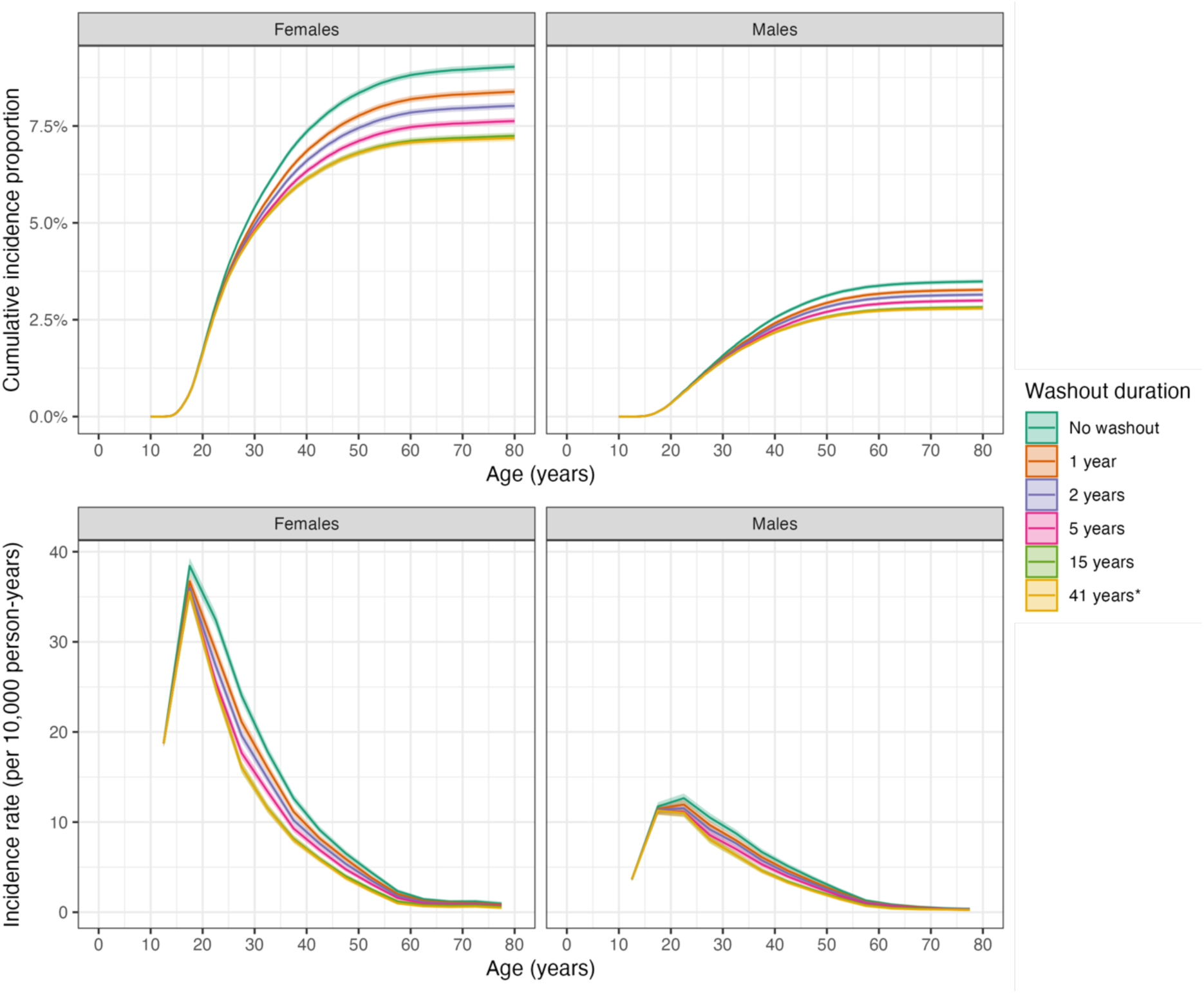
Sex– and age-specific cumulative incidence proportion and incidence rates of personality disorders in Denmark in 2010-2021 using different durations of washout periods (in the period 1969-2009) to identify and exclude individuals with prevalent diagnoses of personality disorders at start of follow-up.

**eFigure 9.**
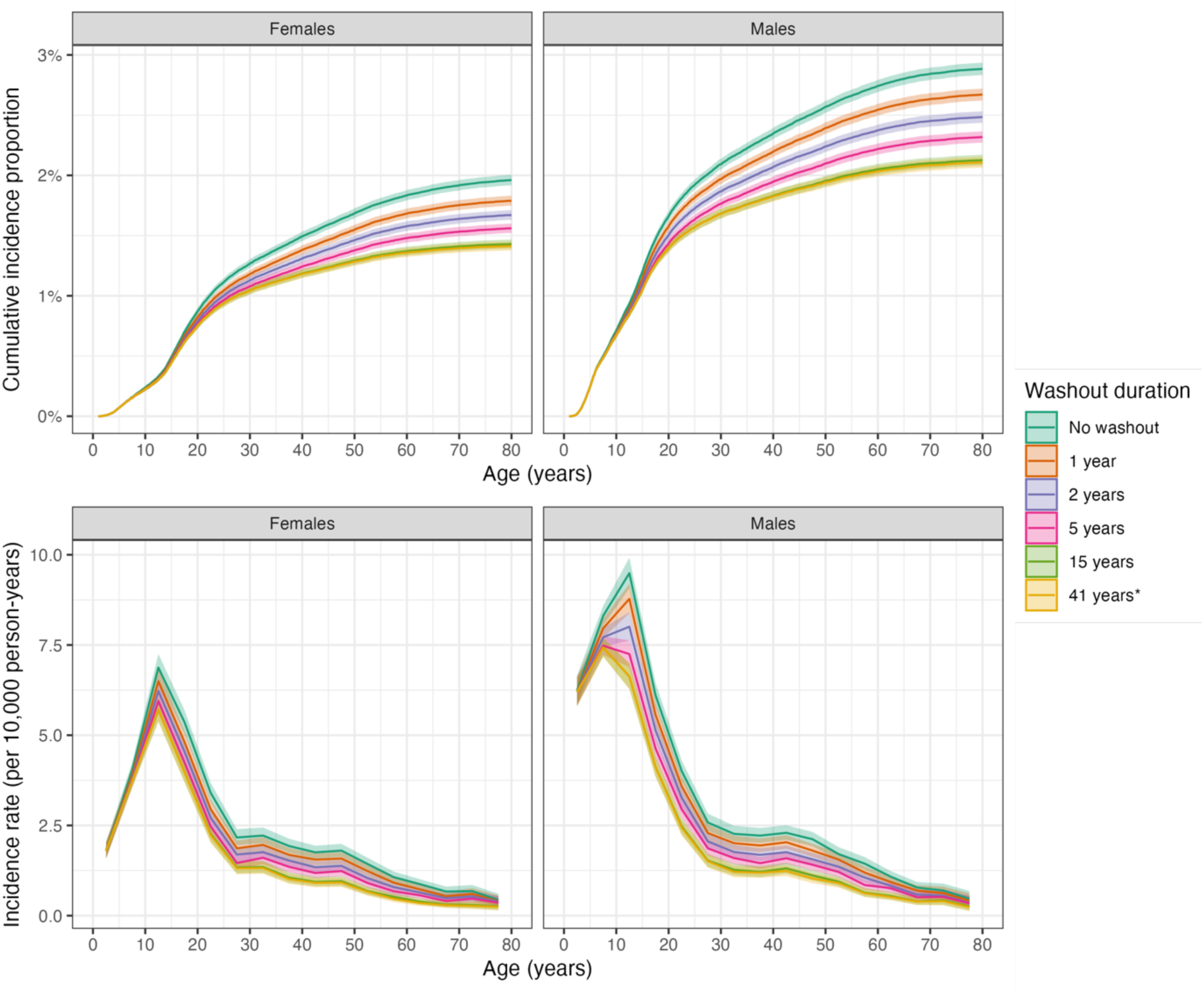
Sex– and age-specific cumulative incidence proportion and incidence rates of intellectual disabilities in Denmark in 2010-2021 using different durations of washout periods (in the period 1969-2009) to identify and exclude individuals with prevalent diagnoses of intellectual disabilities at start of follow-up.

